# Biobank-scale characterization of Alzheimer’s disease and related dementias identifies potential disease-causing variants, risk factors, and genetic modifiers across diverse ancestries

**DOI:** 10.1101/2024.11.03.24313587

**Authors:** Marzieh Khani, Fulya Akçimen, Spencer M. Grant, S. Can Akerman, Paul Suhwan Lee, Faraz Faghri, Hampton Leonard, Jonggeol Jeffrey Kim, Mary B. Makarious, Mathew J. Koretsky, Jeffrey D Rothstein, Cornelis Blauwendraat, Mike A. Nalls, Andrew Singleton, Sara Bandres-Ciga

## Abstract

Alzheimer’s disease and related dementias (AD/ADRDs) pose a significant global public health challenge, underscored by the intricate interplay of genetic and environmental factors that differ across ancestries. To effectively implement equitable, personalized therapeutic interventions on a global scale, it is essential to identify disease-causing mutations and genetic risk and resilience factors across diverse ancestral backgrounds. Exploring genetic-phenotypic correlations across the globe enhances the generalizability of research findings, contributing to a more inclusive and universal understanding of disease. This study leveraged biobank-scale data to conduct the largest multi-ancestry whole-genome sequencing characterization of AD/ADRDs. We aimed to build a valuable catalog of potential disease-causing, genetic risk and resilience variants impacting the etiology of these conditions. We thoroughly characterized genetic variants from key genes associated with AD/ADRDs across 11 genetic ancestries, utilizing data from All of Us, UK Biobank, 100,000 Genomes Project, Alzheimer’s Disease Sequencing Project, and the Accelerating Medicines Partnership in Parkinson’s Disease, including a total of 25,001 cases and 93,542 controls. We prioritized 116 variants possibly linked to disease, including 18 known pathogenic and 98 novel variants. We detected previously described disease-causing variants among controls, leading us to question their pathogenicity. Notably, we showed a higher frequency of *APOE* ε4/ε4 carriers among individuals of African and African Admixed ancestry compared to other ancestries, confirming ancestry-driven modulation of *APOE*-associated AD/ADRDs. A thorough assessment of *APOE* revealed a disease-modifying effect conferred by the *TOMM40*:rs11556505, *APOE*:rs449647, *19q13.31*:rs10423769, *NOCT*:rs13116075, *CASS4*:rs6024870, and *LRRC37A*:rs2732703 variants among *APOE* ε4 carriers across different ancestries. In summary, we compiled the most extensive catalog of established and novel genetic variants in known genes increasing risk or conferring resistance to AD/ADRDs across diverse ancestries, providing clinical insights into their genetic-phenotypic correlations. The findings from this investigation hold significant implications for potential clinical trials and therapeutic interventions on a global scale. Finally, we present an accessible and user-friendly platform for the AD/ADRDs research community to help inform and support basic, translational, and clinical research on these debilitating conditions (https://niacard.shinyapps.io/MAMBARD_browser/).

## Introduction

In 2023, the World Health Organization reported that dementia affects approximately 55 million people worldwide [1]. This number is expected to reach approximately 152.8 million (ranging from 130.8 to 175.9 million) by 2050 [2], placing a significant burden on healthcare infrastructure. Alzheimer’s disease (AD), the most common form of dementia, represents roughly 60-70% of all cases [1]. Less prevalent forms, such as dementia with Lewy bodies (DLB) and frontotemporal dementia (FTD), each account for 10–15% of dementia cases [3,4].

Most of the research conducted thus far on the genetic underpinnings of dementia has primarily focused on populations of European ancestry, limiting the generalizability of findings [5]. Growing evidence indicates significant differences in the genetic architecture of disease among diverse ancestral populations, which raises concerns about the development of therapeutic interventions based on genetic targets primarily identified in a single population. Expanding research to include diverse ancestries is crucial for precision therapeutics. In the new era of personalized medicine, achieving accurate and effective disease-modifying treatments requires a comprehensive understanding of these diseases in a global context.

In recent years, researchers and healthcare institutions worldwide have undertaken ambitious efforts to create large-scale datasets encompassing diverse genetic ancestries, providing valuable insights into the genetic, environmental, and clinical factors influencing disease susceptibility and progression [6,7]. While more work remains in collecting diverse genetic datasets that are dementia-specific, existing efforts can provide valuable insights into dementia research. Currently, All of Us (AoU), UK Biobank (UKB), 100,000 Genomes Project (100KGP), Alzheimer’s Disease Sequencing Project (ADSP), and the Accelerating Medicines Partnership in Parkinson’s Disease (AMP PD) represent the largest and most prominent publicly available dementia datasets worldwide.

A priority in elucidating the etiology of AD and related dementias (AD/ADRDs) lies in defining cumulative risk; however, very little is known about genetic factors that enhance resistance to or protect against dementia. In genetics, protective variants reduce the risk of developing dementia or delay its onset. They confer protection via a loss-of-function or gain-of-function mechanism and can influence various biological pathways associated with the disease. Resilience variants (disease-modifying factors reducing the penetrance of risk loci) influence the development and course of the disease in individuals already at risk, potentially delaying symptom onset or reducing disease severity by interacting with pre-existing risk variants (genetic modifiers). To the best of our knowledge, 11 protective and 10 resilience variants have been reported in AD, with a particular focus on the role of genetic variation modulating AD risk among homozygous or heterozygous *APOE* ε4 carriers [8–14]. Understanding factors that confer protection or resilience can inform therapeutic strategies to reduce the overall burden of dementia, potentially decreasing healthcare costs and the societal impact of the disease.

In this study, we aimed to conduct the largest and most comprehensive multi-ancestry whole-genome sequencing characterization of AD/ADRDs potential disease-causing variation, as well as risk, protective, and disease-modifying factors leveraging biobank-scale data. We screened genetic variants in key genes linked to these conditions, including *APP*, *PSEN1*, *PSEN2*, *TREM2*, *MAPT*, *GRN*, *GBA*, *SNCA*, and *APOE* across a total of 25,001 AD/ADRD cases and 93,542 control individuals, collectively representing 11 ancestries. Furthermore, we assessed protective and disease-modifying variants among different *APOE* genotype carriers in those ancestry groups. This research is particularly relevant in the context of population-specific target prioritization for therapeutic interventions. Such advancements are crucial, as drug mechanisms supported by genetic insights have a 2.6 times higher likelihood of success than those without such support, underscoring the importance of including diverse genetic data to enhance therapeutic outcomes [15]. Here, we present genetic-phenotypic correlations among identified variants across all datasets and develop a user-friendly platform for the scientific community to help inform and support basic, translational, and clinical research on these debilitating conditions (https://niacard.shinyapps.io/MAMBARD_browser/).

## Methods

Demographic information, including age and sex, was provided in the self-reported survey in AoU and through the UKB, ADSP, and AMP PD portals. Self-reported demographic data of participants in 100KGP were obtained from Data Release V18 (12/21/2023) using the LabKey application incorporated into the research environment. **Figure 1** displays the demographic characteristics of cohorts under study. **Figure 2** shows a summary of our workflow, which we explain in further detail below.

**Figure 1.**
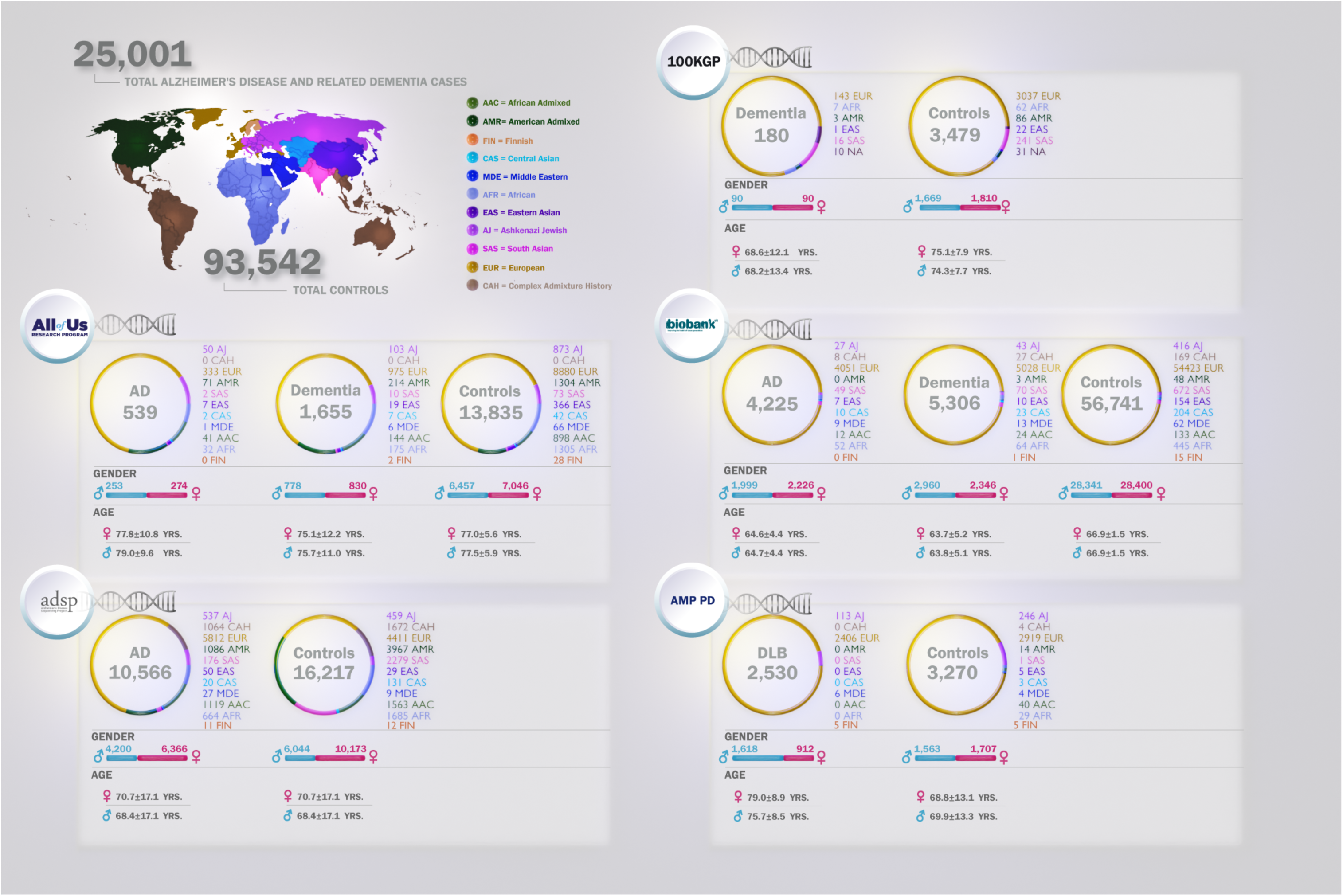
Demographic and clinical characteristics of biobank-scale cohorts under study. The figure illustrates distributions of age, sex, and the number of cases and controls per ancestry across five datasets in this study: All of Us (AoU), Alzheimer’s Disease Sequencing Project (ADSP), 100,000 Genomes Project (100KGP), UK Biobank (UKB), and Accelerating Medicines Partnership in Parkinson’s Disease (AMP PD). Ancestries represented include European (EUR), African (AFR), American Admixed (AMR), African Admixed (AAC), Ashkenazi Jewish (AJ), Central Asian (CAS), Eastern Asian (EAS), South Asian (SAS), Middle Eastern (MDE), Finnish (FIN), and Complex Admixture History (CAH).

**Figure 2.**
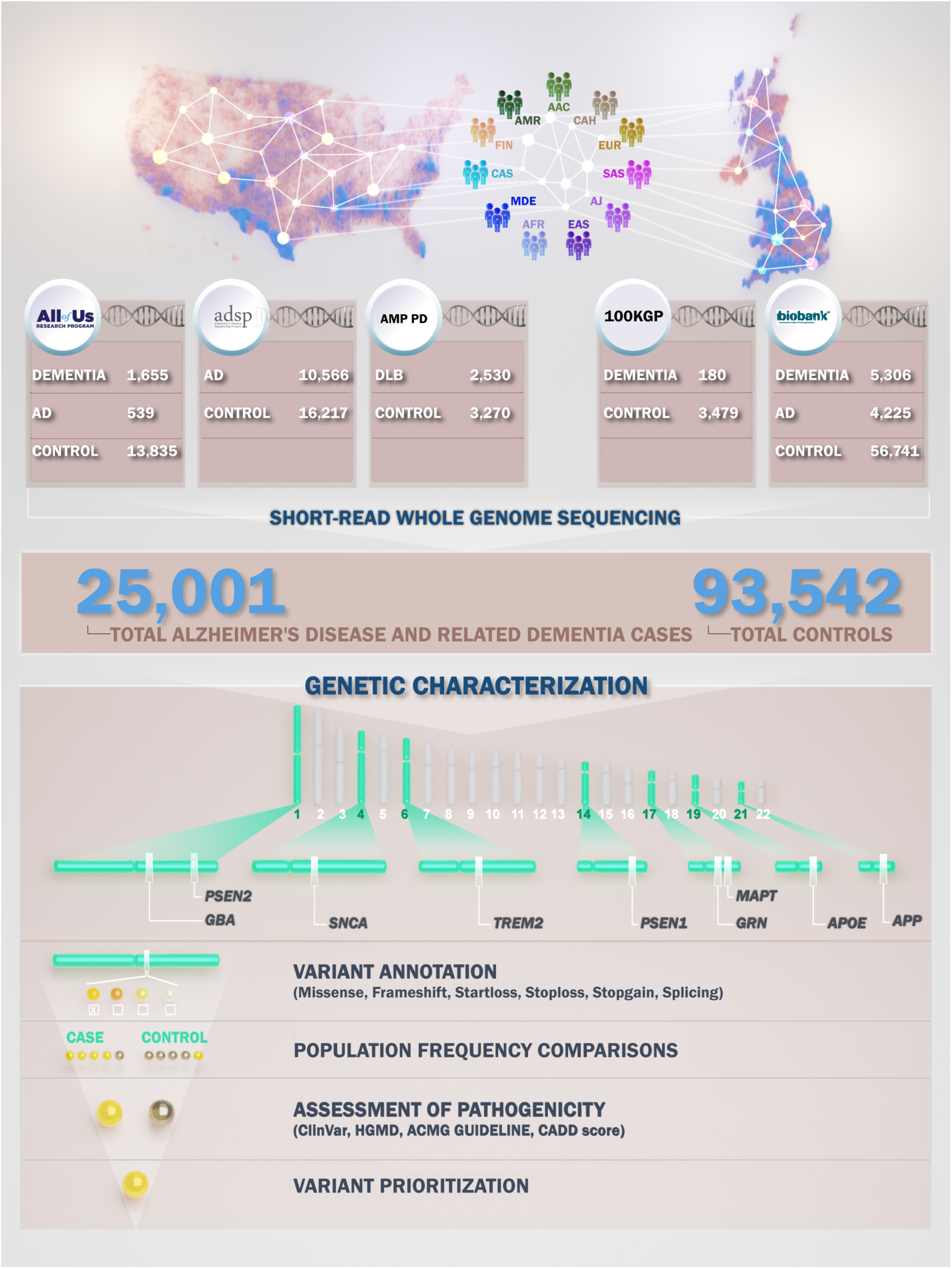
Workflow. Our workflow begins with creating cohorts within the datasets. We leverage short-read whole genome sequencing data to characterize genes of interest. Variant annotation focuses on missense, frameshift, start loss, stop loss, stop gain, and splicing variants. Next, we compare the frequency of identified variants in cases and controls. Pathogenicity assessment involves using ClinVar, Human Gene Mutation Database (HGMD), American College of Medical Genetics and Genomics (ACMG) guidelines, and Combined Annotation Dependent Depletion (CADD) scores. Finally, we prioritize variants that are present only in the case cohort and have a CADD score greater than 20.

### Discovery phase: All of Us

The All of Us (AoU) Research Program (allofus.nih.gov) launched by the United States National Institutes of Health (NIH) endeavors to enhance precision health strategies by assembling rich longitudinal data from over one million diverse participants in the United States. The program emphasizes health equity by engaging underrepresented groups in biomedical research. This biobank includes a wide range of health information, including genetic, lifestyle data, and electronic health records (EHRs), among others, making it a valuable resource for studying the genetic and environmental determinants of various diseases, including AD/ADRDs [6,16].

### Data Acquisition

We accessed the AoU data through the AoU researcher workbench cloud computing environment (https://workbench.researchallofus.org/), utilizing Python and R programming languages for querying. We used the online AoU data browser (https://databrowser.researchallofus.org/variants) to extract genetic variants from short-read whole genome sequencing (WGS) data. The selected variants were filtered for protein-altering or splicing mechanisms for further analysis.

### Cohort Creation

We generated WGS cohorts using the cohort-creating tool in the AoU Researcher Workbench. AD/ADRD cases were selected based on the condition domain in the EHRs. Controls were selected among individuals ≥ 65 years old without any neurological condition in their EHRs, family history, or neurological history in their self-reported surveys. In total, 539 AD cases, 1,655 related dementias, and 13,835 controls were included in the study.

### Whole genome sequencing protocol and quality control assessment

WGS was conducted by the Genome Centers funded by the AoU Research Program [6,17], all of which followed the same protocols. Sequencing details are described elsewhere [18]. Phenotypic data, ancestry features, and principal components (PCs) were generated using Hail within the AoU Researcher Workbench (https://support.researchallofus.org/hc/en-us/articles/4614687617556-How-the-All-of-Us-Genomic-data-are-organized). Ancestry annotation and relatedness were determined using the PC-relate method in Hail and duplicated samples and one of each related participant pair with KINSHIP less than 0.1 being excluded from the Hail data [19] (https://support.researchallofus.org/hc/en-us/articles/4614687617556-How-the-All-of-Us-Genomic-data-are-organized). Flagged individuals and low-quality variants (qc.call_rate < 0.90) were removed from the analysis.

### Variant Filtering and Analysis

We utilized protein-altering and splicing variants (‘WGS_EXOME_SPLIT_HAIL_PATH’) for our analysis, obtained following the tutorial ‘How to Work with AoU Genomic Data (Hail - Plink) (v7).’ The largest intervals for genomic positions were obtained from the UCSC Genome Browser (https://genome.ucsc.edu/).

Variant datasets were obtained as described in the related workspace (see “How to Work with AoU Genomic Data (Hail - Plink) (v7)” for further details). Genomic positions (GRCh38) for each gene were extracted from the Hail variant dataset. Variant-level quality assessments were applied as described in the Manipulate Hail Variant Dataset tutorial (see the “How to Work with AoU Genomic Data” workspace for further details). VCF files containing the cohorts in the current study were generated using BCFtools v1.12 [20]. Allele frequency and zygosity of each resulting variant were calculated per ancestry using PLINK v2.0 [21] in each of the AD, related dementias, and control cohorts.

#### Discovery phase: UK Biobank

The UK Biobank (UKB) (https://www.ukbiobank.ac.uk/) is a large-scale biomedical dataset containing detailed genetic, clinical, and lifestyle information from over 500,000 participants aged 40 to 69 years in the United Kingdom. Each participant’s profile includes a diverse array of phenotypic and health-related information. Additionally, the participants’ health has been followed long-term, primarily through linkage to a wide range of health-related records, enabling the validation and characterization of health-related outcomes [7]. This dataset has been instrumental in advancing research on various health conditions, including AD/ADRDs, by facilitating large-scale genome-wide association studies and rare, deleterious variant analyses [7].

### Cohort Creation

We accessed UKB data (https://www.ukbiobank.ac.uk/) through the DNAnexus cloud computing environment, utilizing the Python programming language for querying. Three experimental cohorts were defined: AD, related dementias, and controls. The AD cohort was defined by the UKB field ID 42020, using diagnoses according to the UKB’s algorithmically defined outcomes v2.0 (https://biobank.ndph.ox.ac.uk/ukb/refer.cgi?id=460). The related dementia cohort was defined by the UKB field ID 42018, using the UKB’s algorithmically defined “Dementia” classification, with the added step of excluding any individuals in the aforementioned AD cohort. The control cohort includes individuals ≥ 65 years without any neurological condition or family history of neurological disorders. Relatedness was calculated with KING [22], and individuals closer than cousins were removed by KINSHIP > 0.0884 to ensure no pair of participants across all three cohorts were related. In total, 4,225 AD cases, 5,306 related dementias, and 56,741 controls were included in the study.

### Whole genome sequencing protocol and quality control assessment

Sequencing was conducted using the NovaSeq 6000 platform [23]. These data were then analyzed with the DRAGEN v3.7.8 (Illumina, San Diego, CA, USA) software. Alignment was performed against the GRCh38 reference genome. Further details on quality control metrics can be found at https://biobank.ndph.ox.ac.uk/showcase/label.cgi?id=187.

### Data Acquisition

WGS data are stored in the UKB as multi-sample aggregated pVCF files, each representing distinct 20 kbp segments for all participants. Genomic ranges were defined for each gene of interest using Ensembl (https://useast.ensembl.org/index.html). Those pVCF files containing any variants within these genomic ranges were included for analysis. Left alignment and normalization were performed on each of these variants using BCFtools v1.15.1 [24]. Then, ANNOVAR [25] was used to annotate the normalized variants.

### Variant Filtering and Analysis

We filtered variants to include only those within our genes of interest, annotated as either protein-altering or splicing variants and present in any AD and/or related dementia cases. Allele frequency and zygosity of each resulting variant were calculated per ancestry using PLINK v2.0 [21] in each of the AD, related dementias, and control cohorts.

#### Discovery phase: 100,000 Genomes Project

The 100,000 Genomes Project (100KGP) (https://www.genomicsengland.co.uk/) has sequenced and analyzed genomes from over 75,000 participants with rare diseases and family members. Early onset dementia (encompassing FTD) is one of the rare diseases studied by the Neurology and Neurodevelopmental Disorders group within the rare disease domain. Participants were recruited by healthcare professionals and researchers from 13 Genomic Medicine Centres in England. The probands were enrolled in the project if they or their guardian provided written consent for their samples and data to be used in research. Probands and, if feasible, other family members were enrolled according to eligibility criteria set for certain rare disease conditions.

WGS data were utilized from 180 unrelated cases with early-onset dementia (encompassing FTD and prion disease) or Parkinson’s disease (PD) with dementia phenotype and 3,479 unrelated controls ≥ 65 years at the time of the analysis. Sequencing and quality control analyses for the 100KGP were previously described elsewhere [26] (https://re-docs.genomicsengland.co.uk/sample_qc/). Protein-altering or splicing variants were obtained using the Exomiser variant prioritization application [27]. Candidate variants were extracted from a multi-sample aggregated VCF provided in the Genomics England research environment.

#### Replication phase: Alzheimer’s Disease Sequencing Project

The Alzheimer’s Disease Sequencing Project (ADSP) (https://adsp.niagads.org/), supported by the National Institute on Aging and the National Human Genome Research Institute, aims to generate data associated with AD/ADRDs. This dataset includes genetic data from thousands of individuals with and without AD, facilitating the discovery of novel genetic risk factors and pathways underlying the disease.

We used data from the ADSP dataset (v4) for this study, which included a total of 10,566 AD cases and 16,217 controls. The control cohort includes individuals ≥ 65 years without any neurological condition or family history of neurological disorders. Samples were excluded from further analysis if the sample call rate was less than 95%, the genetically determined sex did not match the sex reported in clinical data, or excess heterozygosity was detected (|F| statistics > 0.25). For quality control purposes, an MAF threshold of 0.1% was used. The missingness rate and allele frequency of these variants were calculated for each ancestry using PLINK v2.0 [21] and PLINK v1.9 [28]. Variant quality control included removing variants with Hardy-Weinberg Equilibrium P < 1 × 10^−4^ in control samples, differential missingness by case-control status at P ≤ 1 × 10^−4^, and non-random missingness by haplotype at P ≤ 1 × 10^−4^. Relatedness was calculated with KING [22], and individuals closer than cousins were removed by KINSHIP > 0.0884. Duplicated samples were also removed. ADSP includes a range of cohorts, including extensive family cohorts and cohorts with progressive supranuclear palsy (PSP), corticobasal degeneration, mild cognitive impairment (MCI), and DLB patients. Only samples labeled as definite AD or control were included in this analysis. We meticulously screened for identified genetic variants with a CADD score > 20 that were present across any of the three discovery datasets (AoU, 100KGP, and UKB) in the ADSP cohort.

#### Replication phase: Accelerating Medicines Partnership in Parkinson’s Disease

Accelerating Medicines Partnership (AMP) (https://fnih.org/our-programs/accelerating-medicines-partnership-amp/) is a public-private initiative that aims to transform the current model for developing new diagnostics and treatments by jointly identifying and validating promising biological targets for therapeutics. It was launched in 2014 by the NIH, the U.S. Food and Drug Administration, multiple biopharmaceutical and life science companies, and several non-profit organizations. AMP PD focuses on advancing research into PD-related disorders and leverages cutting-edge technologies and large-scale data analysis to identify key genetic variants, biomarkers, and therapeutic targets associated with PD-related disorders, with the ultimate goal of developing novel treatments and improving patient outcomes.

We used AMP PD Release 3 genomic data, focusing specifically on DLB cases and controls. Samples were excluded from further analysis if the sample call rate was less than 95%, the genetically determined sex did not match the sex reported in clinical data, or excess heterozygosity was detected (|F| statistics > 0.25). Variant quality control included removing variants with missingness above 0.05%. Relatedness was calculated with KING [22], and individuals closer than first cousins were removed by KINSHIP > 0.0884. After quality control and ancestry prediction, this dataset contains a total of 2,530 DLB cases and 3,270 controls, characterized as individuals ≥ 65 years without any neurological condition or family history of any neurological disorders. We screened for identified variants with a CADD score > 20 that were present across all three discovery datasets (AoU, 100KGP, and UKB) within AMP PD. Allele frequency of these variants per ancestry was calculated using PLINK v2.0 [21] and PLINK v1.9 [28].

#### Ancestry Prediction Analysis

All samples in AoU, UKB, ADSP, and AMP PD datasets underwent a custom ancestry prediction pipeline included in the GenoTools package (https://github.com/dvitale199/GenoTools) [29]. In brief, ancestry was defined using reference panels from the 1000 Genomes Project, the Human Genome Diversity Project, and an Ashkenazi Jewish population dataset. We used a panel of 4,008 samples from 1000 Genomes Project and the Gene Expression Omnibus database (www.ncbi.nlm.nih.gov/geo; accession no. GSE23636) to define ancestry reference populations. The reference panel was then reduced to exclude palindromic SNPs (AT or TA or GC or CG). SNPs with minor allele frequency (MAF) < 0.05, genotyping call rate < 0.99, and HWE P < 1E-4 in the reference panel were further excluded. Variants overlapping between the reference panel SNP set and the samples of interest were then extracted. Any missing genotypes were imputed using the mean of that particular variant in the reference panel. The reference panel samples were split into an 80/20 train/test set, and then PCs were fitted using the set of overlapping SNPs described previously. The PCs were then transformed via UMAP to represent global genetic population substructure and stochastic variation. A classifier was then trained on these UMAP transformations of the PCs (linear support vector). Based on the test data from the reference panel and at 5-fold cross-validation, 11 ancestries were predicted consistently with balanced accuracies greater than 0.95.

Genetic ancestry in 100KGP was estimated by generating PCs for 1000 Genomes Project phase 3 samples and projecting all participants onto the super populations in the 1000 Genomes Project, as described elsewhere (https://re-docs.genomicsengland.co.uk/ancestry_inference/). Despite our efforts to utilize GenoTools, we encountered significant challenges during its implementation in Genomics England’s High-Performance Computing Cluster (HPC). Consequently, GenoTools and Genomics England’s HPC were incompatible in this context. PCA plots across all biobanks are shown in **Supplementary Figure 1**.

#### Evaluation of potential disease-causing mutations, risk factors, and disease risk modifiers across ancestries

In the discovery phase, variants were filtered out based on their presence in control individuals across biobanks. To prioritize potential disease-causing mutations, we followed the American College of Medical Genetics and Genomics (ACMG) guidelines (https://wintervar.wglab.org/), leveraging existing clinical and population databases and pathogenicity predictors including the Human Gene Mutation Database (HGMD) (https://www.hgmd.cf.ac.uk/ac/index.php), dbSNP (https://www.ncbi.nlm.nih.gov/snp/), gnomAD (https://gnomad.broadinstitute.org/), ClinVar (https://www.ncbi.nlm.nih.gov/clinvar/), PolyPhen-2 (http://genetics.bwh.harvard.edu/pph2/), and Combined Annotation Dependent Depletion (CADD) scores (GRCh38-v1.7) (https://cadd.gs.washington.edu/).

Secondly, we investigated *APOE*, the major risk factor for AD/ADRDs, across diverse ancestries. We used PLINK (v1.9 and v2.0) [21,28] to extract genotypes for two *APOE* variants, rs429358 (chr19:44908684-44908685) and rs7412 (chr19:44908821-44908823), as a proxy for *APOE* allele status (ε1, ε2, ε3, and ε4) in the AoU, UKB, ADSP, and AMP PD datasets. Data analysis was conducted as reported elsewhere (https://github.com/neurogenetics/APOE_genotypes). In the 100KGP dataset, *APOE* genotypes were analyzed using PLINK v2.0 [21] in a multi-sample aggregated VCF provided in the Genomics England research environment. Subsequently, we calculated the number of individuals with each genotype per ancestry and their frequency percentages.

Finally, we assessed disease modifiers for *APOE* ε4 homozygous and heterozygous carriers specifically. A total of 21 variants, previously identified as either protective (n=11) or resilient (n=10), were extracted from all datasets using the same protocol previously described. Among them, *ABCA7*:rs72973581-A, *APP*:rs466433-G, *APP*:rs364048-C, *NOCT*:rs13116075-G, *SORL1*:rs11218343-C, *SLC24A4*:rs12881735-C, *CASS4*:rs6024870-A, *EPHA1*:rs11762262-A, *SPPL2A*:rs59685680-G, *APP*:rs63750847-T, *PLCG2*:rs72824905-G are protective, while *19q13.31*:rs10423769-A, *APOE*:rs449647-T, *FN1*:rs140926439-T, *FN1*:rs116558455-A, *RELN*:rs201731543-C, *TOMM40*:rs11556505-T, *RAB10*:rs142787485-G, *LRRC37A*:rs2732703-G, *NFIC*:rs9749589-A, and the *APOE3* Christchurch:rs121918393-A variant are reported to be resilient. These variants were then checked across all *APOE* genotypes and ancestries. Carrier frequencies (either heterozygous or homozygous) were calculated for each *APOE* genotype and ancestry and were then combined across each of the datasets. In AoU, a variant dataset in Hail format (WGS_VDS_PATH) was used for the analysis. PLINK v2.0 [21] and R v4.3.1 (https://www.r-project.org/) were used to assess the protective model (which evaluates the effect of each protective/disease-modifying variant on the phenotype), conditional model (which evaluates the effect of each protective/disease-modifying variant on the phenotype in the presence of *APOE* (ε4, ε4/ε4, ε3/ε3)), R² model (which evaluates the correlation of each protective/disease-modifying variant with *APOE* (ε4, ε4/ε4, ε3/ε3)), and interaction model (which evaluates putative interactions between each protective/disease-modifying variant and *APOE* (ε4, ε4/ε4, ε3/ε3) on the phenotype). Logistic and linear regression analyses, adjusting for *APOE* status, sex, age, and PCs, were applied in the most well-powered dataset (ADSP) to explore these effects.

## Results

### Large-scale genetic characterization nominates known and novel potential disease-causing variants associated with Alzheimer’s disease and related dementias

A summary of the identified variants can be found in **Figure 3**. We identified a total of 159 variants in the *APP, PSEN1, PSEN2, TREM2, GRN, MAPT, GBA1*, and *SNCA* genes within the AoU dataset. All variants and their allele frequencies across different ancestries are available in **Supplementary Table 1**. Among these, 30 genetic variants were present only in cases and had a CADD score > 20 (CADD score > 20 means that the variant is among the top 1% most pathogenic in the genome, as a proxy for its deleteriousness). All 30 of these identified variants were heterozygous. Of these, four were previously reported in AD or FTD (**Table 1**), while 26 were novel (**Table 2**). Of the four known variants, two were found in cases of European ancestry, one of African ancestry, and one of American Admixed ancestry. Among the 26 novel variants, 18 were found in cases of European ancestry, three in cases of African ancestry, two in cases of American ancestry, one in a case of Ashkenazi Jewish ancestry, and two in cases of African Admixed ancestry.

**Figure 3.**
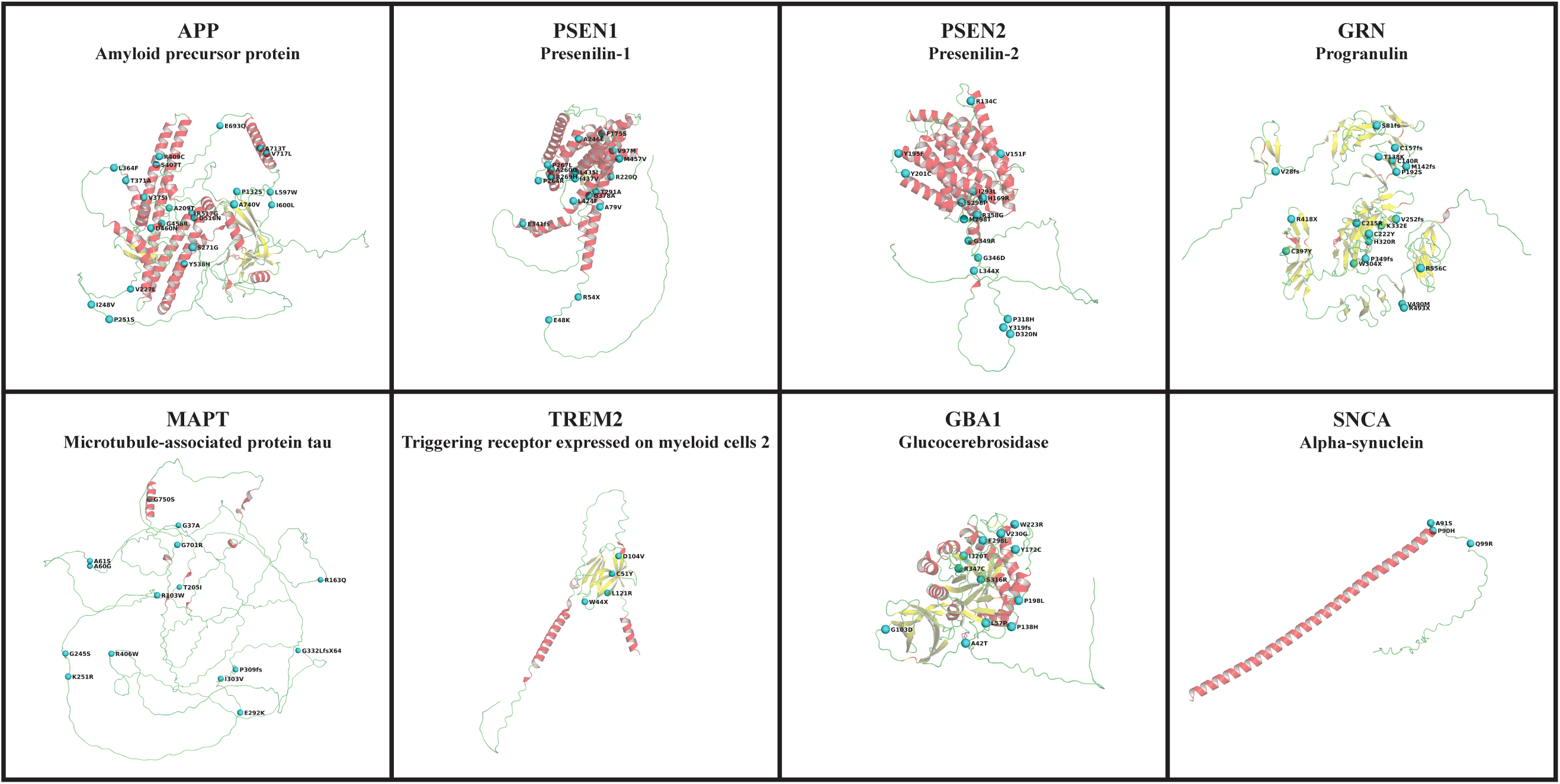
Mutation sites from identified genetic variants mapped on the predicted protein structures encoded by genes associated with AD/ADRDs. The predicted protein structures encoded by eight genes associated with AD/ADRDs (*APP*, *PSEN1*, *PSEN2*, *TREM2*, *GBA1*, *GRN*, *MAPT*, and *SNCA*) were obtained from the EMBL AlphaFold Protein Structure Database to ensure that all of the residues are present in each protein structure. PyMOL v. 2.6.0 was used to represent the protein structures and their associated mutation sites from identified genetic variants. The yellow color shows beta sheets, the red color shows alpha helices, and the green color shows connecting loops and turns.

**Table 1.**
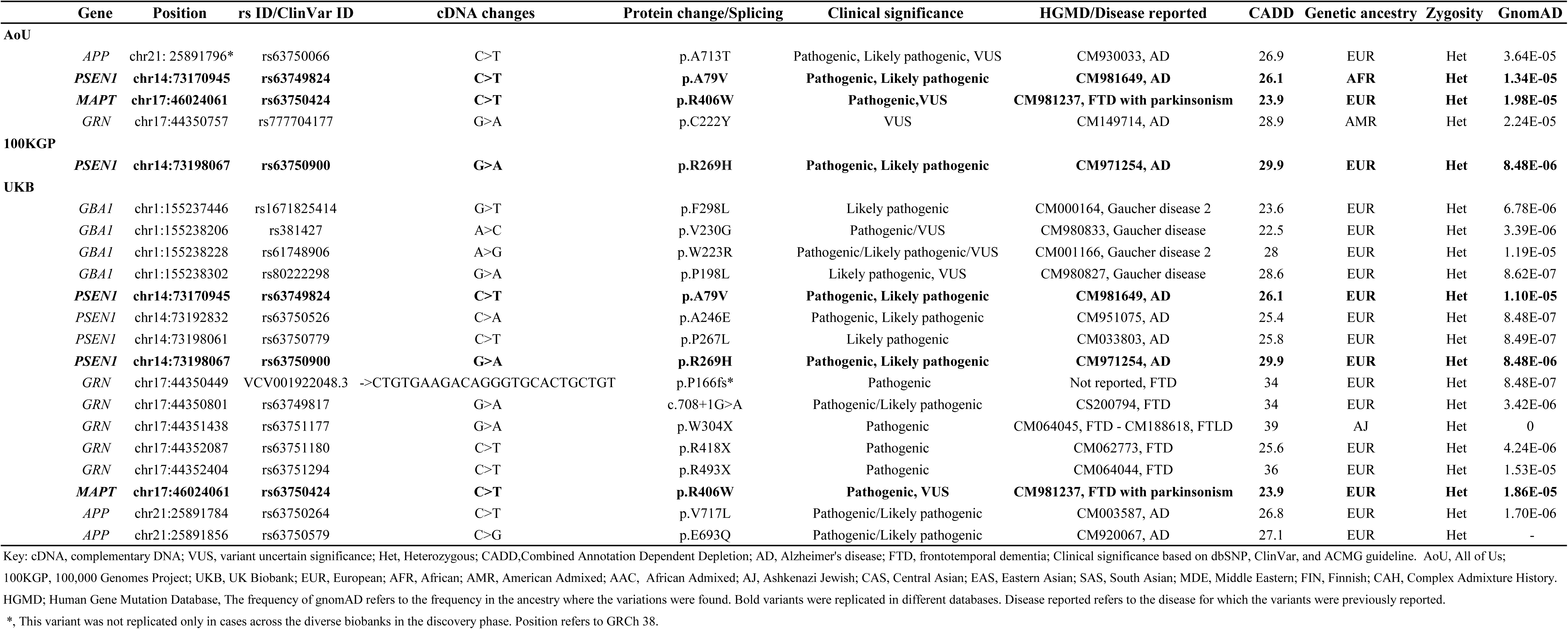
Discovery phase: Multi-ancestry summary of known potential disease-causing variants only present in Alzheimer’s disease and related dementia cases in AoU, 100KGP and UKB.

**Table 2.**
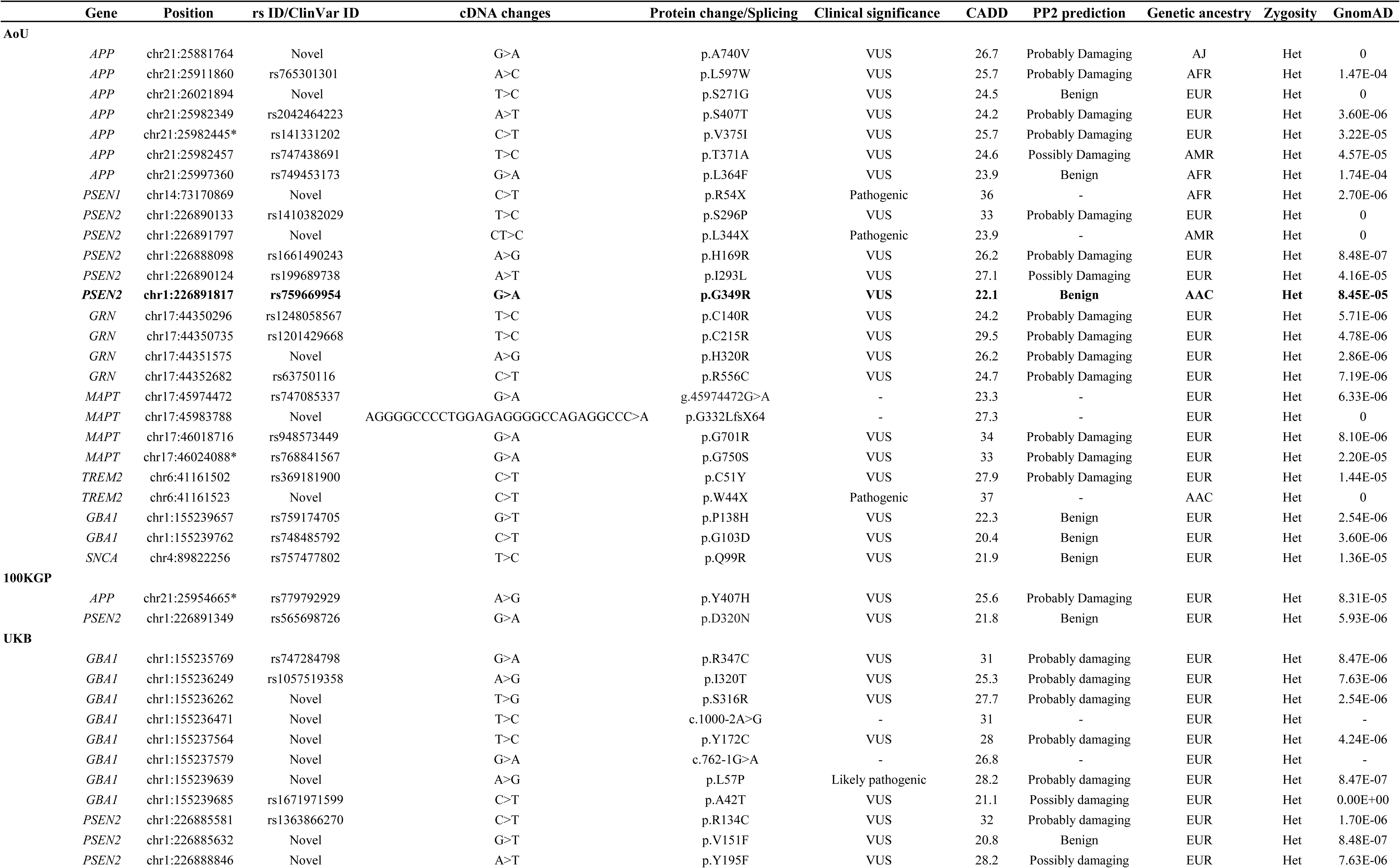
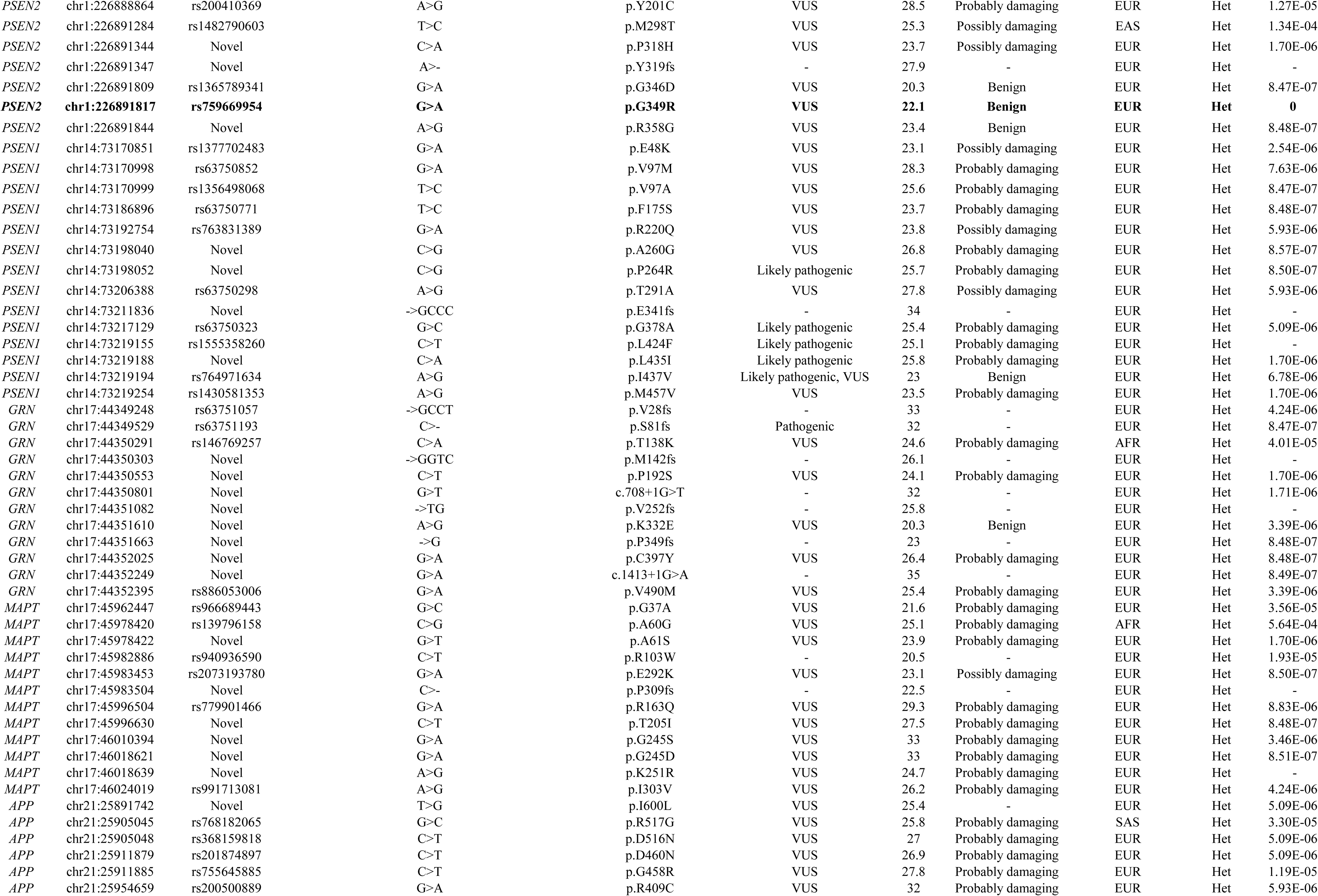
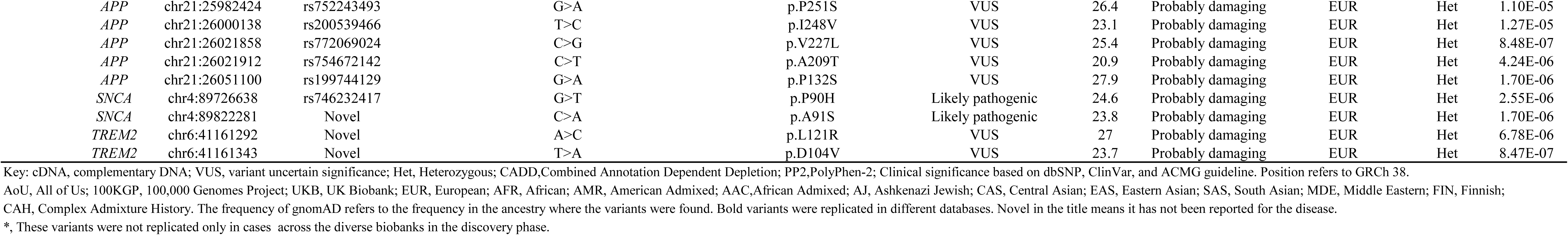
Discovery phase: Multi-ancestry summary of novel potential disease-causing variants only present in Alzheimer’s disease and related dementia cases in AoU, 100KGP and UKB.

Within the UKB, we identified a total of 650 variants in the *APP, PSEN1, PSEN2, TREM2, GRN, MAPT, GBA1,* and *SNCA* genes (**Supplementary Table 2**). Among these, 87 variants were present only in cases and had a CADD score > 20. All 87 identified variants were heterozygous. Of these, 16 were previously reported as disease-causing in AD, FTD, frontotemporal lobar degeneration (FTLD), and Gaucher disease (**Table 1**), while 71 were novel (**Table 2**). A majority (n = 82) of the variants were identified in individuals of European genetic ancestry, two in cases of African ancestry, and one each in cases of South Asian, East Asian, and Ashkenazi Jewish ancestries, respectively. The allele frequencies of the variants across different ancestries are reported in **Supplementary Table 2**.

We identified a total of 11 variants in the *APP*, *PSEN1*, *PSEN2*, *GRN*, and *GBA1* genes within the 100KGP data (**Supplementary Table 3**). Among cases, no variants were identified in the *MAPT*, *TREM2*, and *SNCA* genes. Of the 11 variants, three were only present in cases and had a CADD score > 20. All three identified variants were heterozygous and previously reported in individuals of European ancestry. Among these three variants, *PSEN1* p.R269H had been previously reported as a cause of AD **(Table 1)**, while the remaining two variants in the *APP* and *PSEN2* genes were novel (**Table 2)**. The allele frequency of each variant is presented in **Supplementary Table 3**.

#### Replication analyses support the relevance of identified genetic variation across diverse ancestries

Six variants identified in AoU, 16 identified in UKB, and three identified in 100KGP were replicated in AD cases in the ADSP cohort **(Table 3)**. Among the six variants found in AoU that were replicated in ADSP, two variants — *APP* p.A713T and *PSEN1* p.A79V — had been previously reported, while four variants — *APP* p.L597W, *MAPT* p.G701R, *MAPT* p.G750S, and *SNCA* p.Q99R — were novel, with *APP* p.L597W being found in African, African Admixed, and Complex Admixture History ancestries. Searching for other dementia cases resulted in the identification of *APP* p.A713T in one DLB case and *PSEN1* p.A79V in a possible AD case according to ADSP diagnosis criteria. The allele frequency of each variant per genetic ancestry in cases and controls is reported in **Table 3**.

**Table 3.**
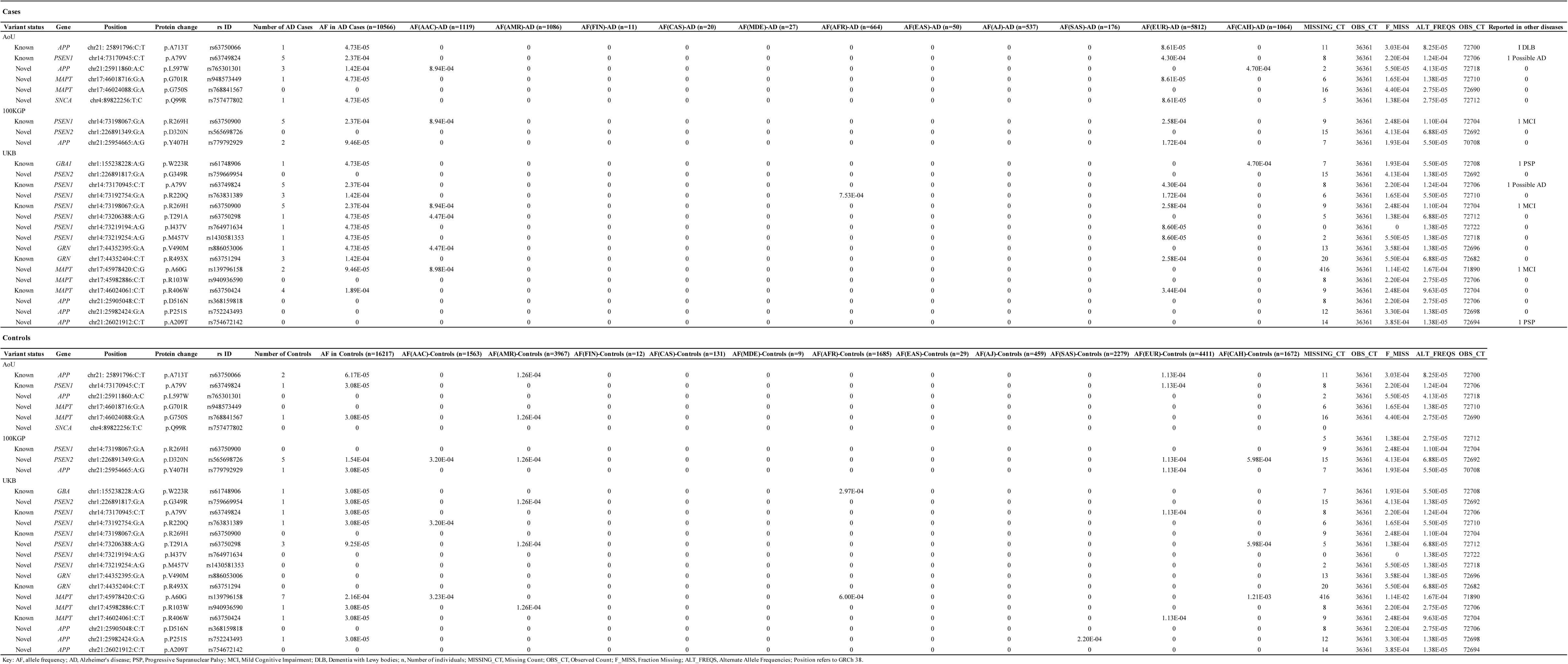
Replication phase: potential disease-causing variants only present in Alzheimer’s disease and related dementia cases in ADSP.

Among the three variants identified in the 100KGP dataset that were replicated in the ADSP cohort, *PSEN1* p.R269H was previously reported while *PSEN2* p.D320N and *APP* p.Y407H were novel. We observed the previously reported *PSEN1* p.R269H variant in five cases and no control participants. This variant was found in European ancestry individuals from the 100KGP cohort and was also observed in individuals of African Admixed ancestry (two cases) and European ancestry (three cases) in the ADSP dataset. Of the novel variants, the *PSEN2* p.D320N variant was found in five controls and was not observed in any cases, while the *APP* p.Y407H variant was observed in two cases and one control. Searching for additional cases led to the discovery of *PSEN1* p.R269H in a patient with MCI.

Among the 16 variants identified in the UKB cohort that were replicated in the ADSP dataset, five variants — *PSEN1* p.A79V (five cases and one control), *PSEN1* p.R269H (five cases), *GRN* p.R493X (three cases), *MAPT* p.R406W (four cases and one control), and *GBA1* p.W223R (one case and one control) — have been previously reported. The remaining 11 variants were novel. Most of the novel variants were found in European cases in the UKB. The *PSEN1* p.R269H variant was found in cases of both African Admixed and European ancestries, and *GBA1* p.W223R was found in a case of Complex Admixture History ancestry and a control of African ancestry. The three remaining known variants were observed in individuals of European ancestry in the ADSP cohort. Novel variants identified in non-European participants include *PSEN1* p.R220Q (one African case, two European cases, and one African Admixed control), *PSEN1* p.T291A (one African Admixed case, one American Admixed control, and two controls with Complex Admixture History), *MAPT* p.A60G (two African Admixed cases, one African Admixed control, two African controls, and four controls with Complex Admixture History), *GRN* p.V490M (one African Admixed case), *MAPT* p.R103W (one American Admixed control), and *APP* p.P251S (one South Asian control). Searching for other cases resulted in the identification of *PSEN1* p.A79V in one possible AD patient, *PSEN1* p.R269H and *MAPT* p.A60G in two independent MCI patients, and *GBA1* p.W223R and *APP* p.A209T in two independent PSP patients **(Table 3)**.

We identified a novel *SNCA* variant (p.Q99R) in the AoU dataset, while the UKB dataset revealed two additional variants in *SNCA*: p.P90H and p.A91S. Both p.P90H and p.A91S were predicted to be likely pathogenic according to prediction estimates and have not been previously reported as disease-causing. Notably, the *SNCA* p.Q99R variant was replicated in the ADSP cohort. All three variants were heterozygous, and none of these variants were found in any controls across these datasets. However, the age at onset of these variant carriers is not consistent with a potential disease-causing deleterious effect.

Our analyses of multiple datasets identified 11 variants (three from AoU, seven from UKB, and one from 100KGP) that were absent in the ADSP control cohort.

Across each of our discovery datasets, we identified five candidate variants — *APP* p.A713T, *MAPT* p.G750S, *GRN* p.V490M, *GRN* p.R493X, and *APP* p.D516N — present in AMP PD. *GRN* p.V490M was present in one control and no cases, *GRN* p.R493X was present in one case and no controls, while the other three were present across both cases and controls. The allele frequencies of these variants are detailed in **Table 4**.

**Table 4.**
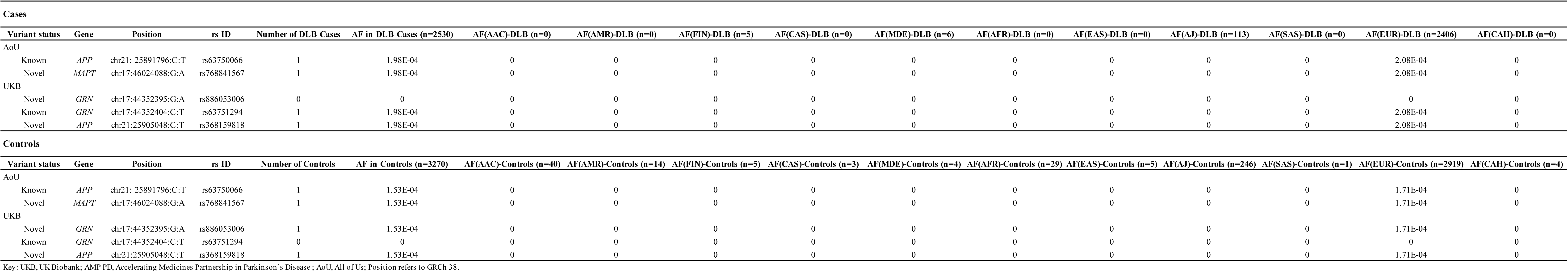
Replication phase: potential disease-causing variants only present in Alzheimer’s disease and related dementia cases in AMP PD.

Among the 116 variants identified in this study, 13 were found exclusively in non-European ancestries. Notably, *APP*:p.L597W and *MAPT*:p.A60G were replicated in African and African Admixed ancestries across different datasets. These data highlight the potential significance of these variants in groups that are often underrepresented in genomic studies.

**Supplementary Figure 2** shows the allele frequencies of all identified known and novel variants with CADD > 20 in the discovery and replication phases across all ancestries in each biobank.

#### Previously reported disease-causing variants raise questions about potential pathogenicity

Although the *SNCA* p.H50Q variant was initially identified as a pathogenic mutation in PD [30], subsequent research has challenged its pathogenicity [31]. Our study confirms that it is not pathogenic across other synucleinopathies such as DLB, based on its occurrence in five European controls in AoU and 28 European controls in UKB.

Additionally, several research studies have reported the *APP* p.A713T variant to be disease-causing [32,33]. In our study, we found this variant in heterozygous state in five control individuals: two in UKB, two in ADSP, and one in AMP PD. Interestingly, the *APP* p.E665D variant, which has been widely reported to cause AD [34,35], was found in one control in AoU in her late 70s. However, it is possible that the variant shows incomplete penetrance, or that this individual may harbor unidentified resilient genetic variation. Another previous study evaluating the role of *APP* p.E665D questioned the pathogenicity of this variant [36].

*GBA1* coding variants in heterozygous state generally exhibit incomplete penetrance and act as genetic risk factors. Homozygous *GBA1* variants, including the p.T75del and c.115+1G>A mutations have been reported to cause Gaucher disease [37–40]. We found these two variants in a heterozygous state in one case and one control in AoU. *GBA1* p.T75del was found in individuals of African ancestry, and *GBA1* c.115+1G>A was found in individuals of European ancestry in both a case and a control. The *GBA1* c.115+1G>A variant was also found in nine European controls in the UKB cohort. Thirteen additional heterozygous variants in *GBA1* — p.R502C, p.A495P, p.L483R, p.D448H, p.E427X, p.G416S, p.N409S, p.R398X, p.R296Q, p.G241R, p.N227S, p.S212X, and p.R159W — were identified in our study, and have been reported as disease-causing for Gaucher disease in homozygous state. Three variants in *GRN*, including two loss of function variants (including p.Q130fs and p.Y294X) and one splicing variant (c.708+6_708+9del), have been previously reported to cause FTD, FTLD, and neurodegenerative disease [37,41–55]. Each of these 16 variants were found in several control individuals **(Supplementary Table 4)**.

#### Genetic-phenotypic correlations provide valuable clinical insights

Clinical data for the identified variants are summarized in **Supplementary Tables 5 and 6**. Here, we briefly explain the main findings.

The *GRN* p.R493X variant is this gene’s most reported pathogenic mutation. This variant has been associated with several types of dementia, including FTD, FTLD, primary progressive aphasia, AD, and corticobasal degeneration. It is known to be more frequently identified among FTD cases, particularly in early-onset forms [56]. In one study investigating the genetics underlying disease etiology in 1,118 DLB patients, this variant was reported in a single case, presenting a wide range of neurological phenotypes that could not lead to a conclusive diagnosis. Severe dementia, parkinsonism, and visual hallucinations suggested a clinical diagnosis of AD or mixed vascular dementia. However, the final neuropathological diagnosis was suggested to be AD, DLB, and argyrophilic grain disease [57]. We identified this variant in four European AD patients, three of whom presented with early onset in their fifties. Interestingly, we also identified this variant in a DLB patient in her early 60s. Neuropathological data and McKeith criteria [58] strongly supported a diagnosis of DLB in this patient. Although this variant has been widely reported across different types of dementia, our finding is the first report of this variant in DLB with a McKeith criteria of “high likelihood of DLB,” expanding the etiological spectrum of *GRN* variation **(Supplementary Table 5)**.

*GRN* p.C222Y was previously reported in a familial AD case from Latin American (Caribbean Hispanic) ancestry [59,60]. While the AAO for this patient was not reported, the mean AAO for the cohorts under study was 56.9 years (SD = 7.29), with a range between 40–73 years. In our study, we identified this variant in an individual of American Admixed ancestry with dementia in his late 40s and a disease duration of 11 years to date. This finding reinforces the role of this variant in early-onset disease.

There are several other interesting findings regarding variants in *GRN*. The *GRN* c.708+1G>A variant was previously reported in several FTD, FTLD, and corticobasal syndrome (CBS) cases, mostly early-onset [55,61]. We identified this variant in two European AD cases, both diagnosed in their 70s, marking the first report of this variant in late-onset Alzheimer’s disease (LOAD). The *GRN* p.P166fsX variant was previously reported in an early-onset behavioral variant FTD case [62]. In our study, we identified this variant in a European dementia case diagnosed in her mid 70s with a disease duration of 8 years to date. The *GRN* p.R418X variant is identified in the literature in two cases of FTLD with ubiquitin-positive inclusions (FTLD-U) with an AAO of 49 and 60 years [63]. We identified this variant in a European dementia case in her early 70s. Both findings represent the first report of these variants in late-onset dementia.

*PSEN1* R269H is a known pathogenic variant causing early-onset Alzheimer’s disease (EOAD) [64,65]. However, it has been previously reported in only two LOAD cases [66,67]. In our study, we identified this variant in European and African Admixed ancestries in a total of 12 cases (eight AD and four related dementias), six of which were early-onset (≤65 years) and six were late-onset (>65 years). This finding underscores the potential for *PSEN1* p.R269H to contribute to LOAD with reduced penetrance. Additionally, one EOAD case that presented with hallucinations [68] and another that manifested a behavioral presentation [69] have been reported to carry this variant. In this study, we identified *PSEN1* p.R269H in one FTD patient in the 100KGP cohort, marking the first report of this variant in FTD.

*MAPT* p.R406W has been reported in several familial cases of FTD with parkinsonism, all with early onset [70]. There are only two articles related to this variant in AD. The first describes a family with AD-like symptoms, with an average AAO of 61 years [71], and the other reports a familial AD case with an AAO of 50 years [72]. In our study, we identified this variant in nine AD cases, with a mean AAO of 61 years. This finding underscores the role of this variant in EOAD.

Several variants in *GBA1*, such as p.F298L, p.V230G, p.W223R, and p.P198L, have been previously reported in Gaucher disease patients. In our study, we identified these variants in heterozygous state in one AD case and five dementia cases, all with late onset. *GBA1* p.W223R was found in one AD case of Complex Admixture History ancestry. *GBA1* mutations are known to confer an increased risk for dementia in PD and DLB. Notably, they have not been previously suggested to contribute to AD.

Similarly, *APP* p.E693Q has been reported in a few AD cases. In our study, we identified it in a related dementia case and no controls. This finding suggests that this variant may also be implicated in other types of dementia.

Several known variants identified in this study confirm previous findings related to disease type and onset. For example, the *APP* p.V717L variant has been reported in numerous AD cases, primarily in early-onset forms [73,74]. In our study, we identified this variant in two cases of EOAD with AAO ranges of 51-55 years and 56-60 years, respectively. Additional examples are reported in **Supplementary Tables 5 and 6**.

In AoU, the *SNCA* p.Q99R variant was found in a female patient diagnosed in her late 60s with unspecified dementia without behavioral disturbance. In ADSP, the variant was identified in a male patient diagnosed with pure AD in his early 70s. *SNCA* p.P90H and p.A91S were found in two males in their late 70s in the UKB cohort. All four patients were of European ancestry. Previously reported mutations in *SNCA* are known to cause early-onset PD and DLB [75,76]. The mean AAO in patients carrying *SNCA* mutations in this study is 72.75 years. These data suggest that these variants may not be disease-causing but could represent rare risk factors despite their absence in controls and the replication of p.Q99R across datasets.

Novel variants found in this study that may potentially be associated with early-onset dementia include: p.L597W, p.V375I, p.L364F, p.A209T, p.D460N, p.R409C, and p.V227L variants in *APP;* p.R54X and p.M457V in *PSEN1*; p.H169R, p.D320N, and p.G349R in *PSEN2;* p.R556C and p.V28fs in *GRN*; p.G332LfsX64 and p.G701R in *MAPT*; p.G103D and p.A42T in *GBA1;* and *TREM2* p.W44X. Among these variants, *APP* p.L364F and *PSEN1* p.R54X were found in vascular dementia cases with AAO ranges of 41-45 years and 46-50 years, respectively. Additionally, *PSEN2* p.D320N was found in an FTD case with an AAO in his mid-50s. Another notable finding is the identification of *GRN* p.R556C in a dementia case with an AAO in her mid-30s.

#### *APOE* drives different population-attributable risk for Alzheimer’s disease and related dementias

The summary of our findings on ancestry-specific effects of *APOE* on AD/ADRDs is depicted in **Figure 4**, **Table 5**, and **Supplementary Figure 3**. In AoU, UKB, and 100KGP, the *APOE* ε4/ε4 genotype exhibits a higher frequency among both AD patients and control individuals of African and African Admixed ancestries compared to Europeans. Related dementia patients show similar results in UKB. In AoU, related dementia cases of African Admixed ancestry show a higher frequency than Europeans, while frequencies are similar between Africans and Europeans, likely due to the limited number of individuals with African ancestry in this dataset. In ADSP, the frequencies of this genotype among AD patients are similar across the three ancestries. Among control individuals in ADSP, *APOE* ε4/ε4 is more frequent in African Admixed and African ancestries than in Europeans, as previously reported [77]. Notably, the *APOE* ε4/ε4 genotype was absent from African and African Admixed DLB cases and controls in the AMP PD dataset. The frequency of *APOE* ε4/ε4 in Europeans was higher in cases compared to controls in AMP PD.

**Figure 4.**
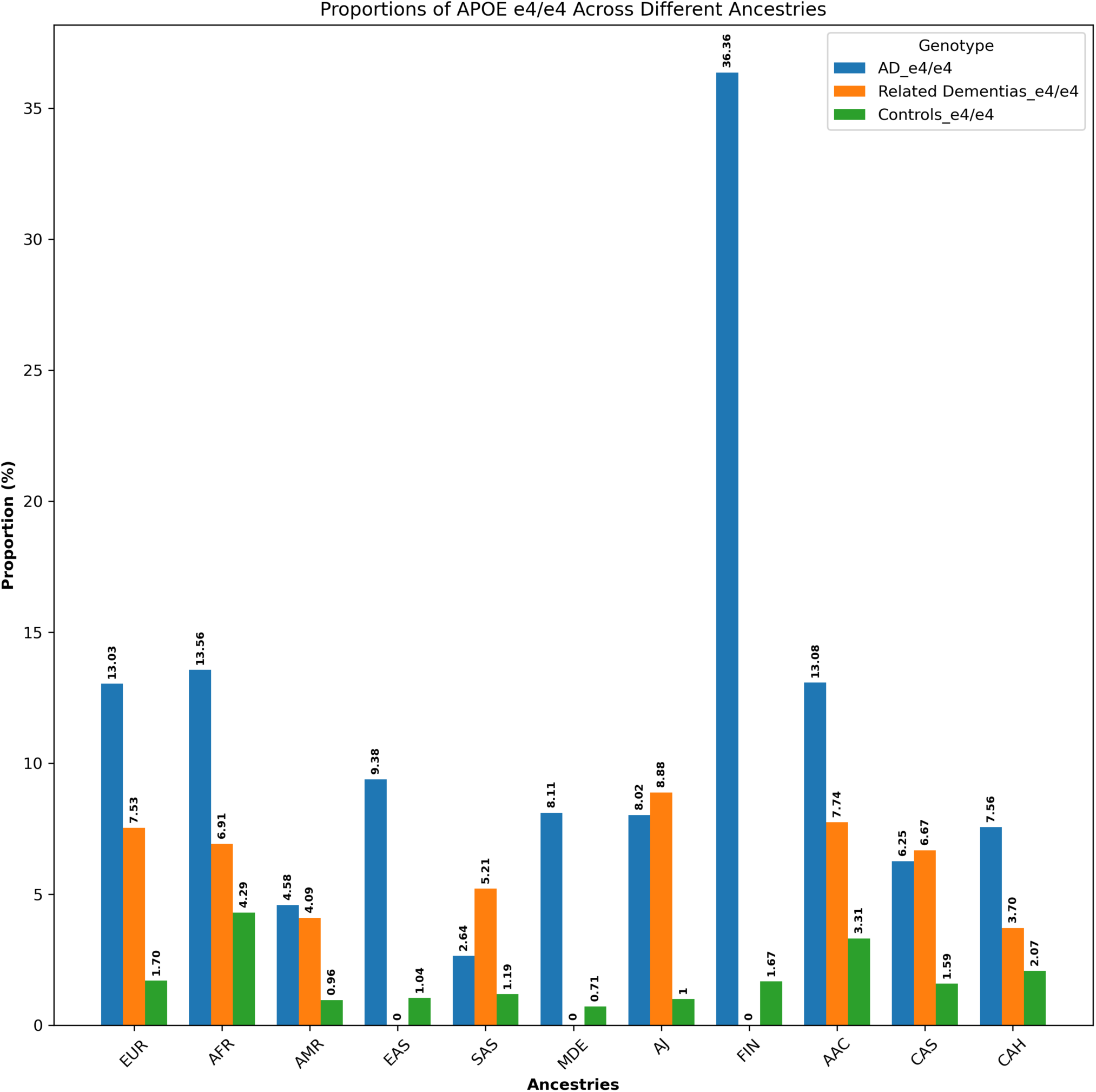
Proportions of *APOE* ε4/ε4 across 11 genetic ancestries in Alzheimer’s disease, related dementias, and controls in all datasets.

**Table 5.** Multi-ancestry summary of *APOE* genotypes in Alzheimer’s disease and related dementia cases and controls in AoU, ADSP, UKB, AMP PD and 100KGP.

| AoU |  |  |  |  |  |  |  |  |  |  |  |  |
| --- | --- | --- | --- | --- | --- | --- | --- | --- | --- | --- | --- | --- |
| AD |  |  |  |  |  |  |  |  |  |  |  |  |
| Genotypes | Number of participants (n, %) | EUR (n, %) | AFR (n, %) | AMR (n, %) | EAS (n, %) | SAS (n, %) | MDE (n, %) | AJ (n, %) | FIN (n, %) | AAC (n, %) | CAS (n, %) | CAH (n, %) |
| ε1/ε1 | 0, 0% | 0, 0% | 0, 0% | 0, 0% | 0, 0% | 0, 0% | 0, 0% | 0, 0% | 0, 0% | 0, 0% | 0, 0% | 0, 0% |
| ε1/ε2 | 0, 0% | 0, 0% | 0, 0% | 0, 0% | 0, 0% | 0, 0% | 0, 0% | 0, 0% | 0, 0% | 0, 0% | 0, 0% | 0, 0% |
| ε1/ε4 | 0, 0% | 0, 0% | 0, 0% | 0, 0% | 0, 0% | 0, 0% | 0, 0% | 0, 0% | 0, 0% | 0, 0% | 0, 0% | 0, 0% |
| ε2/ε2 | 3, 0.55% | 2, 0.6% | 1, 3.12% | 0, 0% | 0, 0% | 0, 0% | 0, 0% | 0, 0% | 0, 0% | 0, 0% | 0, 0% | 0, 0% |
| ε2/ε3 | 46, 8.53% | 26, 7.80% | 2, 6.25% | 4, 5.63% | 2, 28.57% | 0, 0% | 0, 0% | 5, 10% | 0, 0% | 7, 17.07% | 0, 0% | 0, 0% |
| ε2/ε4 or ε1/ε3 | 11, 2.04% | 5, 1.50% | 2, 6.25% | 1, 1.40% | 0, 0% | 0, 0% | 0, 0% | 1, 2% | 0, 0% | 2, 4.87% | 0, 0% | 0, 0% |
| ε3/ε3 | 281, 52.13% | 171, 51.35% | 13, 40.62% | 46, 64.78% | 2, 28.57% | 1, 50% | 1, 100% | 26, 52% | 0, 0% | 20, 48.78% | 1, 50% | 0, 0% |
| ε3/ε4 | 158, 29.31% | 106, 31.83% | 9, 28.12% | 19, 26.76% | 3, 42.85% | 0, 0% | 0, 0% | 11, 22% | 0, 0% | 9, 21.95% | 1, 50% | 0, 0% |
| ε4/ε4 | 40, 7.42% | 23, 6.90% | 5, 15.62% | 1, 1.40% | 0, 0% | 1, 50% | 0, 0% | 7, 14% | 0, 0% | 3, 7.31% | 0, 0% | 0, 0% |
| Total | 539 | 333 | 32 | 71 | 7 | 2 | 1 | 50 | 0 | 41 | 2 | 0 |
| Dementia |  |  |  |  |  |  |  |  |  |  |  |  |
| Genotypes | Number of participants (n, %) | EUR (n, %) | AFR (n, %) | AMR (n, %) | EAS (n, %) | SAS (n, %) | MDE (n, %) | AJ (n, %) | FIN (n, %) | AAC (n, %) | CAS (n, %) | CAH (n, %) |
| 00_CC, unknown | 1, 0.06% | 1, 0.09% | 0, 0% | 0, 0% | 0, 0% | 0, 0% | 0, 0% | 0, 0% | 0, 0% | 0, 0% | 0, 0% | 0, 0% |
| ε1/ε1 | 0, 0% | 0, 0% | 0, 0% | 0, 0% | 0, 0% | 0, 0% | 0, 0% | 0, 0% | 0, 0% | 0, 0% | 0, 0% | 0, 0% |
| ε1/ε2 | 0, 0% | 0, 0% | 0, 0% | 0, 0% | 0, 0% | 0, 0% | 0, 0% | 0, 0% | 0, 0% | 0, 0% | 0, 0% | 0, 0% |
| ε1/ε4 | 0, 0% | 0, 0% | 0, 0% | 0, 0% | 0, 0% | 0, 0% | 0, 0% | 0, 0% | 0, 0% | 0, 0% | 0, 0% | 0, 0% |
| ε2/ε2 | 12, 0.72% | 7, 0.71% | 1, 0.57% | 2, 0.93% | 0, 0% | 0, 0% | 0, 0% | 1, 0.97% | 0, 0% | 1, 0.69% | 0, 0% | 0, 0% |
| ε2/ε3 | 162, 9.78% | 88, 9.02% | 24, 13.71% | 19, 8.87% | 4, 21.05% | 1, 10% | 0, 0% | 8, 7.76% | 0, 0% | 18, 12.5% | 0, 0% | 0, 0% |
| ε2/ε4 or ε1/ε3 | 38, 2.29% | 17, 1.74% | 12, 6.85% | 1, 0.46% | 0, 0% | 0, 0% | 0, 0% | 2, 1.94% | 0, 0% | 6, 4.16% | 0, 0% | 0, 0% |
| ε3/ε3 | 912, 55.10% | 551, 56.51% | 77, 44% | 130, 60.74% | 10, 52.63% | 7, 70% | 6, 100% | 60, 58.25% | 2, 100% | 66, 45.83% | 3, 42.85% | 0, 0% |
| ε3/ε4 | 449, 27.12% | 266, 27.28% | 53, 30.28% | 54, 25.23% | 5, 26.31% | 1, 10% | 0, 0% | 22, 21.35% | 0, 0% | 45, 31.25% | 3, 42.85% | 0, 0% |
| ε4/ε4 | 81, 4.89% | 45, 4.61% | 8, 4.57% | 8, 3.73% | 0, 0% | 1, 10% | 0, 0% | 10, 9.70% | 0, 0% | 8, 5.55% | 1, 14.28% | 0, 0% |
| Total | 1655 | 975 | 175 | 214 | 19 | 10 | 6 | 103 | 2 | 144 | 7 | 0 |
| Controls |  |  |  |  |  |  |  |  |  |  |  |  |
| Genotypes | Number of participants (n, %) | EUR (n, %) | AFR (n, %) | AMR (n, %) | EAS (n, %) | SAS (n, %) | MDE (n, %) | AJ (n, %) | FIN (n, %) | AAC (n, %) | CAS (n, %) | CAH (n, %) |
| CT_00, unknown | 1, 0.007% | 1, 0.01% | 0, 0% | 0, 0% | 0, 0% | 0, 0% | 0, 0% | 0, 0% | 0, 0% | 0, 0% | 0, 0% | 0, 0% |
| 00_CC, unknown | 1, 0.007% | 1, 0.01% | 0, 0% | 0, 0% | 0, 0% | 0, 0% | 0, 0% | 0, 0% | 0, 0% | 0, 0% | 0, 0% | 0, 0% |
| ε1/ε1 | 0, 0% | 0, 0% | 0, 0% | 0, 0% | 0, 0% | 0, 0% | 0, 0% | 0, 0% | 0, 0% | 0, 0% | 0, 0% | 0, 0% |
| ε1/ε2 | 0, 0% | 0, 0% | 0, 0% | 0, 0% | 0, 0% | 0, 0% | 0, 0% | 0, 0% | 0, 0% | 0, 0% | 0, 0% | 0, 0% |
| ε1/ε4 | 0, 0% | 0, 0% | 0, 0% | 0, 0% | 0, 0% | 0, 0% | 0, 0% | 0, 0% | 0, 0% | 0, 0% | 0, 0% | 0, 0% |
| ε2/ε2 | 90, 0.65% | 51, 0.57% | 18, 1.37% | 4, 0.3% | 3, 0.81% | 0, 0% | 0, 0% | 6, 0.68% | 0, 0% | 8, 0.89% | 0, 0% | 0, 0% |
| ε2/ε3 | 1734, 12.53% | 1124, 12.65% | 220, 16.85% | 82, 6.28% | 61, 16.66% | 5, 6.84% | 13, 19.69% | 113, 12.94% | 3, 10.71% | 108, 12.02% | 5, 11.90% | 0, 0% |
| ε2/ε4 or ε1/ε3 | 295, 2.13% | 168, 1.89% | 66, 5.05% | 11, 0.84% | 1, 0.27% | 1, 1.36% | 2, 3.03% | 10, 1.14% | 0, 0% | 36, 4.00% | 0, 0% | 0, 0% |
| ε3/ε3 | 8537, 61.70% | 5597, 63.02% | 587, 44.98% | 927, 71.08% | 252, 68.85% | 51, 69.86% | 46, 69.69% | 575, 65.86% | 20, 71.42% | 453, 50.44% | 29, 69.04% | 0, 0% |
| ε3/ε4 | 2930, 21.17% | 1817, 20.46% | 349, 26.74% | 265, 20.32% | 48, 13.11% | 16, 21.91% | 5, 7.57% | 160, 18.32% | 4, 14.28% | 259, 28.84% | 7, 16.66% | 0, 0% |
| ε4/ε4 | 247, 1.78% | 121, 1.36% | 65, 4.98% | 15, 1.15% | 1, 0.27% | 0, 0% | 0, 0% | 9, 1.03% | 1, 3.57% | 34, 3.78% | 1, 2.38% | 0, 0% |
| Total | 13835 | 8880 | 1305 | 1304 | 366 | 73 | 66 | 873 | 28 | 898 | 42 | 0 |
| ADSP |  |  |  |  |  |  |  |  |  |  |  |  |
| AD |  |  |  |  |  |  |  |  |  |  |  |  |
| Genotypes | Number of participants (n, %) | EUR (n, %) | AFR (n, %) | AMR (n, %) | EAS (n, %) | SAS (n, %) | MDE (n, %) | AJ (n, %) | FIN (n, %) | AAC (n, %) | CAS (n, %) | CAH (n, %) |
| 00_00, unknown | 45, 0.42% | 44, 0.75% | 0, 0% | 0, 0% | 0, 0% | 0, 0% | 0, 0% | 1, 0.18% | 0, 0% | 0, 0% | 0, 0% | 0, 0% |
| 00_CC, unknown | 17, 0.16% | 10, 0.17% | 3, 0.45% | 0, 0% | 0, 0% | 0, 0% | 0, 0% | 1, 0.18% | 0, 0% | 2, 0.17% | 0, 0% | 1, 0.09% |
| 00_TC, unknown | 1, 0.00000095% | 0, 0% | 0, 0% | 0, 0% | 0, 0% | 0, 0% | 0, 0% | 1, 0.18% | 0, 0% | 0, 0% | 0, 0% | 0, 0% |
| TT_00, unknown | 7, 0.06% | 7, 0.12% | 0, 0% | 0, 0% | 0, 0% | 0, 0% | 0, 0% | 0, 0% | 0, 0% | 0, 0% | 0, 0% | 0, 0% |
| CT_00, unknown | 22, 0.20% | 22, 0.37% | 0, 0% | 0, 0% | 0, 0% | 0, 0% | 0, 0% | 0, 0% | 0, 0% | 0, 0% | 0, 0% | 0, 0% |
| CC_00, unknown | 9, 0.08% | 9, 0.15% | 0, 0% | 0, 0% | 0, 0% | 0, 0% | 0, 0% | 0, 0% | 0, 0% | 0, 0% | 0, 0% | 0, 0% |
| ε1/ε1 | 0, 0% | 0, 0% | 0, 0% | 0, 0% | 0, 0% | 0, 0% | 0, 0% | 0, 0% | 0, 0% | 0, 0% | 0, 0% | 0, 0% |
| ε1/ε2 | 0, 0% | 0, 0% | 0, 0% | 0, 0% | 0, 0% | 0, 0% | 0, 0% | 0, 0% | 0, 0% | 0, 0% | 0, 0% | 0, 0% |
| ε1/ε4 | 0, 0% | 0, 0% | 0, 0% | 0, 0% | 0, 0% | 0, 0% | 0, 0% | 0, 0% | 0, 0% | 0, 0% | 0, 0% | 0, 0% |
| ε2/ε2 | 21, 0.19% | 7, 0.12% | 4, 0.60% | 0, 0% | 0, 0% | 0, 0% | 0, 0% | 2, 0.37% | 0, 0% | 5, 0.44% | 0, 0% | 3, 0.28% |
| ε2/ε3 | 479, 4.53% | 177, 3.04% | 47, 7.07% | 38, 3.49% | 3, 6% | 15, 8.52% | 1, 3.70% | 18, 3.35% | 0, 0% | 108, 9.65% | 2, 10% | 70, 6.57% |
| ε2/ε4 or ε1/ε3 | 251, 2.37% | 129, 2.21% | 32, 4.81% | 9, 0.82% | 1, 2% | 4, 2.27% | 0, 0% | 11, 2.04% | 0, 0% | 41, 3.66% | 1, 5% | 23, 2.16% |
| ε3/ε3 | 4296, 40.65% | 2070, 35.61% | 210, 31.62% | 686, 63.16% | 24, 48% | 112, 63.63% | 16, 59.25% | 241, 44.87% | 3, 27.27% | 377, 33.69% | 9, 45% | 548, 51.50% |
| ε3/ε4 | 4263, 40.34% | 2596, 44.66% | 285, 42.92% | 301, 27.71% | 16, 32% | 43, 24.43% | 8, 29.62% | 222, 41.34% | 4, 36.36% | 443, 39.58% | 7, 35% | 338, 31.76% |
| ε4/ε4 | 1155, 10.93% | 741, 12.74% | 83, 12.5% | 52, 4, 78% | 6, 12% | 2, 1.13% | 2, 7.40% | 40, 7.44% | 4, 36.36% | 143, 12.77% | 1, 5% | 81, 7.61% |
| Total | 10566 | 5812 | 664 | 1086 | 50 | 176 | 27 | 537 | 11 | 1119 | 20 | 1064 |

| Controls |  |  |  |  |  |  |  |  |  |  |  |  |
| --- | --- | --- | --- | --- | --- | --- | --- | --- | --- | --- | --- | --- |
| Genotypes | Number of participants (n, %) | EUR (n, %) | AFR (n, %) | AMR (n, %) | EAS (n, %) | SAS (n, %) | MDE (n, %) | AJ (n, %) | FIN (n, %) | AAC (n, %) | CAS (n, %) | CAH (n, %) |
| 00_00, unknown | 124, 0.76% | 120, 2.72% | 0, 0% | 0, 0% | 0, 0% | 0, 0% | 0, 0% | 0, 0% | 0, 0% | 1, 0.06% | 0, 0% | 3, 0.17% |
| 00_CC, unknown | 21, 0.12% | 13, 0.29% | 2, 0.11% | 1, 0.02% | 0, 0% | 0, 0% | 0, 0% | 0, 0% | 0, 0% | 4, 0.25% | 0, 0% | 1, 0.05 |
| 00_TC, unknown | 3, 0.01% | 2, 0.04% | 1, 0.05% | 0, 0% | 0, 0% | 0, 0% | 0, 0% | 0, 0% | 0, 0% | 0, 0% | 0, 0% | 0, 0% |
| TT_00, unknown | 5, 0.03% | 3, 0.06% | 0, 0% | 0, 0% | 0, 0% | 0, 0% | 0, 0% | 1, 0.21% | 0, 0% | 0, 0% | 0, 0% | 1, 0.05 |
| CT_00, unknown | 83, 0.51% | 82, 1.85% | 0, 0% | 0, 0% | 0, 0% | 0, 0% | 0, 0% | 1, 0.21% | 0, 0% | 0, 0% | 0, 0% | 0, 0% |
| CC_00, unknown | 8, 0.04% | 8, 0.18% | 0, 0% | 0, 0% | 0, 0% | 0, 0% | 0, 0% | 0, 0% | 0, 0% | 0, 0% | 0, 0% | 0, 0% |
| ε1/ε1 | 0, 0% | 0, 0% | 0, 0% | 0, 0% | 0, 0% | 0, 0% | 0, 0% | 0, 0% | 0, 0% | 0, 0% | 0, 0% | 0, 0% |
| ε1/ε2 | 0, 0% | 0, 0% | 0, 0% | 0, 0% | 0, 0% | 0, 0% | 0, 0% | 0, 0% | 0, 0% | 0, 0% | 0, 0% | 0, 0% |
| ε1/ε4 | 1, 0.006% | 0, 0% | 1, 0.05% | 0, 0% | 0, 0% | 0, 0% | 0, 0% | 0, 0% | 0, 0% | 0, 0% | 0, 0% | 0, 0% |
| ε2/ε2 | 71, 0.43% | 18, 0.40% | 22, 1.30% | 5, 0.12% | 0, 0% | 3, 0.13% | 0, 0% | 1, 0.21% | 0, 0% | 15, 0.95% | 2, 1.52% | 5, 0.29% |
| ε2/ε3 | 1528, 9.42% | 418, 9.47% | 244, 14.48% | 217, 5.47% | 5, 17.24% | 175, 7.67% | 1, 11.11% | 47, 10.23% | 3, 25% | 235, 15.03% | 18, 13.74% | 165, 9.86% |
| ε2/ε4 or ε1/ε3 | 293, 1.80% | 68, 1.54% | 78, 4.62% | 28, 0.70% | 1, 3.44% | 25, 1.09% | 0, 0% | 4, 0.87% | 0, 0% | 61, 3.90% | 1, 0.76% | 27, 1.61% |
| ε3/ε3 | 10197, 62.87% | 2540, 57.58% | 777, 46.11% | 3008, 75.82% | 12, 41.37% | 1649, 72.35% | 6, 66.66% | 270, 58.82% | 5, 41.66% | 765, 48.94% | 86, 65.64% | 1079, 64.53% |
| ε3/ε4 | 3544, 21.85% | 1026, 23.26% | 494, 29.31% | 672, 16.93% | 8, 27.58% | 398, 17.46% | 2, 22.22% | 128, 27.88% | 4, 33.33% | 435, 27.83% | 20, 15.26% | 357, 21.35% |
| ε4/ε4 | 339, 2.09% | 113, 2.56% | 66, 3.91% | 36, 0.90% | 3, 10.34% | 29, 1.27% | 0, 0% | 7, 1.52% | 0, 0% | 47, 3.00% | 4, 3.05% | 34, 2.03% |
| Total | 16217 | 4411 | 1685 | 3967 | 29 | 2279 | 9 | 459 | 12 | 1563 | 131 | 1672 |

| UKB |
| --- |

| AD |  |  |  |  |  |  |  |  |  |  |  |  |
| --- | --- | --- | --- | --- | --- | --- | --- | --- | --- | --- | --- | --- |
| Genotypes | Number of participants (n, %) | EUR (n, %) | AFR (n, %) | AMR (n, %) | EAS (n, %) | SAS (n, %) | MDE (n, %) | AJ (n, %) | FIN (n, %) | AAC (n, %) | CAS (n, %) | CAH (n, %) |
| ε1/ε1 | 0 , 0% | 0 , 0% | 0 , 0% | 0 , 0% | 0 , 0% | 0 , 0% | 0 , 0% | 0 , 0% | 0 , 0% | 0 , 0% | 0 , 0% | 0 , 0% |
| ε1/ε2 | 0 , 0% | 0 , 0% | 0 , 0% | 0 , 0% | 0 , 0% | 0 , 0% | 0 , 0% | 0 , 0% | 0 , 0% | 0 , 0% | 0 , 0% | 0 , 0% |
| ε1/ε4 | 0 , 0% | 0 , 0% | 0 , 0% | 0 , 0% | 0 , 0% | 0 , 0% | 0 , 0% | 0 , 0% | 0 , 0% | 0 , 0% | 0 , 0% | 0 , 0% |
| ε2/ε2 | 9, 0.21% | 8 , 0.20% | 0 , 0% | 0 , 0% | 0 , 0% | 0 , 0% | 1 , 11.11% | 0 , 0% | 0 , 0% | 0 , 0% | 0 , 0% | 0 , 0% |
| ε2/ε3 | 212, 5.02% | 205 , 5.06% | 5 , 9.62% | 0 , 0% | 0 , 0% | 1 , 2.04% | 0 , 0% | 0 , 0% | 0 , 0% | 1 , 8.33% | 0 , 0% | 0 , 0% |
| ε2/ε4 or ε1/ε3 | 102, 0.02% | 97 , 2.39% | 5 , 9.62% | 0 , 0% | 0 , 0% | 0 , 0% | 0 , 0% | 0 , 0% | 0 , 0% | 0 , 0% | 0 , 0% | 0 , 0% |
| ε3/ε3 | 1391, 32.92% | 1321 , 32.61% | 11 , 21.15% | 0 , 0% | 5 , 71.43% | 25 , 51.02% | 6 , 66.67% | 10 , 37.04% | 0 , 0% | 1 , 8.33% | 6 , 60% | 6 , 75% |
| ε3/ε4 | 1931, 45.70% | 1867 , 46.09% | 18 , 34.62% | 0 , 0% | 2 , 28.57% | 20 , 40.82% | 1 , 11.11% | 15 , 55.56% | 0 , 0% | 3 , 25% | 3 , 30% | 2 , 25% |
| ε4/ε4 | 580, 13.73% | 553 , 13.65% | 13 , 25% | 0 , 0% | 0 , 0% | 3 , 6.12% | 1 , 11.11% | 2 , 7.41% | 0 , 0% | 7 , 58.33% | 1 , 10% | 0 , 0% |
| total | 4225 | 4051 | 52 | 0 | 7 | 49 | 9 | 27 | 0 | 12 | 10 | 8 |

| Dementia |  |  |  |  |  |  |  |  |  |  |  |  |
| --- | --- | --- | --- | --- | --- | --- | --- | --- | --- | --- | --- | --- |
| Genotypes | Number of participants (n, %) | EUR (n, %) | AFR (n, %) | AMR (n, %) | EAS (n, %) | SAS (n, %) | MDE (n, %) | AJ (n, %) | FIN (n, %) | AAC (n, %) | CAS (n, %) | CAH (n, %) |
| ε1/ε1 | 0 , 0% | 0 , 0% | 0 , 0% | 0 , 0% | 0 , 0% | 0 , 0% | 0 , 0% | 0 , 0% | 0 , 0% | 0 , 0% | 0 , 0% | 0 , 0% |
| ε1/ε2 | 0 , 0% | 0 , 0% | 0 , 0% | 0 , 0% | 0 , 0% | 0 , 0% | 0 , 0% | 0 , 0% | 0 , 0% | 0 , 0% | 0 , 0% | 0 , 0% |
| ε1/ε4 | 0 , 0% | 0 , 0% | 0 , 0% | 0 , 0% | 0 , 0% | 0 , 0% | 0 , 0% | 0 , 0% | 0 , 0% | 0 , 0% | 0 , 0% | 0 , 0% |
| ε2/ε2 | 24, 0.45% | 23 , 0.46% | 1 , 1.56% | 0 , 0% | 0 , 0% | 0 , 0% | 0 , 0% | 0 , 0% | 0 , 0% | 0 , 0% | 0 , 0% | 0 , 0% |
| ε2/ε3 | 447, 8.42% | 427 , 8.49% | 6 , 9.38% | 0 , 0% | 2 , 20% | 6 , 8.57% | 2 , 15.38% | 2 , 4.65% | 0 , 0% | 1 , 4.17% | 1 , 4.35% | 0 , 0% |
| ε2/ε4 or ε1/ε3 | 165, 3.11% | 153 , 3.04% | 4 , 6.25% | 0 , 0% | 1 , 10% | 1 , 1.43% | 0 , 0% | 1 , 2.33% | 0 , 0% | 3 , 12.50% | 0 , 0% | 2 , 7.41% |
| ε3/ε3 | 2432, 45.83% | 2291 , 45.56% | 23 , 35.94% | 2 , 66.67% | 6 , 60% | 38 , 54.29% | 10 , 76.92% | 26 , 60.47% | 0 , 0% | 8 , 33.33% | 15 , 65.22% | 13 , 48.15% |
| ε3/ε4 | 1832, 34.53% | 1748 , 34.77% | 22 , 34.38% | 1 , 33.33% | 1 , 10% | 21 , 30% | 1 , 7.69% | 13 , 30.23% | 1 , 100% | 7 , 29.17% | 6 , 26.09% | 11 , 40.74% |
| ε4/ε4 | 406, 7.65% | 386 , 7.68% | 8 , 12.50% | 0 , 0% | 0 , 0% | 4 , 5.71% | 0 , 0% | 1 , 2.33% | 0 , 0% | 5 , 20.83% | 1 , 4.35% | 1 , 3.70% |
| total | 5306 | 5028 | 64 | 3 | 10 | 70 | 13 | 43 | 1 | 24 | 23 | 27 |

| Controls |  |  |  |  |  |  |  |  |  |  |  |  |
| --- | --- | --- | --- | --- | --- | --- | --- | --- | --- | --- | --- | --- |
| Genotypes | Number of participants (n, %) | EUR (n, %) | AFR (n, %) | AMR (n, %) | EAS (n, %) | SAS (n, %) | MDE (n, %) | AJ (n, %) | FIN (n, %) | AAC (n, %) | CAS (n, %) | CAH (n, %) |
| ε1/ε1 | 0 , 0% | 0 , 0% | 0 , 0% | 0 , 0% | 0 , 0% | 0 , 0% | 0 , 0% | 0 , 0% | 0 , 0% | 0 , 0% | 0 , 0% | 0 , 0% |
| ε1/ε2 | 0 , 0% | 0 , 0% | 0 , 0% | 0 , 0% | 0 , 0% | 0 , 0% | 0 , 0% | 0 , 0% | 0 , 0% | 0 , 0% | 0 , 0% | 0 , 0% |
| ε1/ε4 | 0 , 0% | 0 , 0% | 0 , 0% | 0 , 0% | 0 , 0% | 0 , 0% | 0 , 0% | 0 , 0% | 0 , 0% | 0 , 0% | 0 , 0% | 0 , 0% |
| ε2/ε2 | 367, 0.65% | 358 , 0.66% | 3 , 0.67% | 0 , 0% | 1 , 0.65% | 1 , 0.15% | 0 , 0% | 2 , 0.48% | 0 , 0% | 0 , 0% | 1 , 0.49% | 1 , 0.59% |
| ε2/ε3 | 7416, 13.07% | 7167 , 13.17% | 67 , 15.06% | 4 , 8.33% | 18 , 11.69% | 51 , 7.59% | 5 , 8.06% | 38 , 9.13% | 3 , 20% | 29 , 21.80% | 16 , 7.84% | 18 , 10.65% |
| ε2/ε4 or ε1/ε3 | 1300, 2.29% | 1231 , 2.26% | 29 , 6.52% | 0 , 0% | 2 , 1.30% | 8 , 1.19% | 0 , 0% | 11 , 2.64% | 0 , 0% | 9 , 6.77% | 4 , 1.96% | 6 , 3.55% |
| ε3/ε3 | 34770, 61.28% | 33277 , 61.15% | 193 , 43.37% | 33 , 68.75% | 103 , 66.88% | 508 , 75.60% | 50 , 80.65% | 284 , 68.27% | 10 , 66.67% | 56 , 42.11% | 153 , 75% | 103 , 60.95% |
| ε3/ε4 | 11946, 21.05% | 11491 , 21.11% | 136 , 30.56% | 11 , 22.92% | 28 , 18.18% | 95 , 14.14% | 6 , 9.68% | 78 , 18.75% | 2 , 13.33% | 33 , 24.81% | 29 , 14.22% | 37 , 21.89% |
| ε4/ε4 | 942, 1.66% | 899 , 1.65% | 17 , 3.82% | 0 , 0% | 2 , 1.30% | 9 , 1.34% | 1 , 1.61% | 3 , 0.72% | 0 , 0% | 6 , 4.51% | 1 , 0.49% | 4 , 2.37% |
| total | 56741 | 54423 | 445 | 48 | 154 | 672 | 62 | 416 | 15 | 133 | 204 | 169 |
| AMP PD |  |  |  |  |  |  |  |  |  |  |  |  |

| DLB |  |  |  |  |  |  |  |  |  |  |  |  |
| --- | --- | --- | --- | --- | --- | --- | --- | --- | --- | --- | --- | --- |
| Genotypes | Number of participants (n, %) | EUR (n, %) | AFR (n, %) | AMR (n, %) | EAS (n, %) | SAS (n, %) | MDE (n, %) | AJ (n, %) | FIN (n, %) | AAC (n, %) | CAS (n, %) | CAH (n, %) |
| unknown,unknown | 0 , 0% | 0 , 0% | 0 , 0% | 0 , 0% | 0 , 0% | 0 , 0% | 0 , 0% | 0 , 0% | 0 , 0% | 0 , 0% | 0 , 0% | 0 , 0% |
| ε1/ε1 | 0 , 0% | 0 , 0% | 0 , 0% | 0 , 0% | 0 , 0% | 0 , 0% | 0 , 0% | 0 , 0% | 0 , 0% | 0 , 0% | 0 , 0% | 0 , 0% |
| ε1/ε2 | 0 , 0% | 0 , 0% | 0 , 0% | 0 , 0% | 0 , 0% | 0 , 0% | 0 , 0% | 0 , 0% | 0 , 0% | 0 , 0% | 0 , 0% | 0 , 0% |
| ε1/ε4 | 0 , 0% | 0 , 0% | 0 , 0% | 0 , 0% | 0 , 0% | 0 , 0% | 0 , 0% | 0 , 0% | 0 , 0% | 0 , 0% | 0 , 0% | 0 , 0% |
| ε2/ε2 | 5, 0.20% | 5 , 0.21% | 0 , 0% | 0 , 0% | 0 , 0% | 0 , 0% | 0 , 0% | 0 , 0% | 0 , 0% | 0 , 0% | 0 , 0% | 0 , 0% |
| ε2/ε3 | 191, 7.55% | 179 , 7.44% | 0 , 0% | 0 , 0% | 0 , 0% | 0 , 0% | 0 , 0% | 12 , 10.62% | 0 , 0% | 0 , 0% | 0 , 0% | 0 , 0% |
| ε2/ε4 or ε1/ε3 | 84, 3.32% | 78 , 3.24% | 0 , 0% | 0 , 0% | 0 , 0% | 0 , 0% | 0 , 0% | 6 , 5.31% | 0 , 0% | 0 , 0% | 0 , 0% | 0 , 0% |
| ε3/ε3 | 1162, 45.93% | 1108 , 46.05% | 0 , 0% | 0 , 0% | 0 , 0% | 0 , 0% | 3 , 50% | 48 , 42.48% | 3 , 60% | 0 , 0% | 0 , 0% | 0 , 0% |
| ε3/ε4 | 882, 34.86% | 842 , 35% | 0 , 0% | 0 , 0% | 0 , 0% | 0 , 0% | 3 , 50% | 35 , 30.97% | 2 , 40% | 0 , 0% | 0 , 0% | 0 , 0% |
| ε4/ε4 | 206, 8.14% | 194 , 8.06% | 0 , 0% | 0 , 0% | 0 , 0% | 0 , 0% | 0 , 0% | 12 , 10.62% | 0 , 0% | 0 , 0% | 0 , 0% | 0 , 0% |
| total | 2530 | 2406 | 0 | 0 | 0 | 0 | 6 | 113 | 5 | 0 | 0 | 0 |

| Controls |  |  |  |  |  |  |  |  |  |  |  |  |
| --- | --- | --- | --- | --- | --- | --- | --- | --- | --- | --- | --- | --- |
| Genotypes | Number of participants (n, %) | EUR (n, %) | AFR (n, %) | AMR (n, %) | EAS (n, %) | SAS (n, %) | MDE (n, %) | AJ (n, %) | FIN (n, %) | AAC (n, %) | CAS (n, %) | CAH (n, %) |
| unknown,unknown | 4, 0.12% | 0 , 0% | 0 , 0% | 0 , 0% | 0 , 0% | 1 , 100% | 0 , 0% | 0 , 0% | 0 , 0% | 0 , 0% | 3 , 100% | 0 , 0% |
| ε1/ε1 | 0 , 0% | 0 , 0% | 0 , 0% | 0 , 0% | 0 , 0% | 0 , 0% | 0 , 0% | 0 , 0% | 0 , 0% | 0 , 0% | 0 , 0% | 0 , 0% |
| ε1/ε2 | 0 , 0% | 0 , 0% | 0 , 0% | 0 , 0% | 0 , 0% | 0 , 0% | 0 , 0% | 0 , 0% | 0 , 0% | 0 , 0% | 0 , 0% | 0 , 0% |
| ε1/ε4 | 0 , 0% | 0 , 0% | 0 , 0% | 0 , 0% | 0 , 0% | 0 , 0% | 0 , 0% | 0 , 0% | 0 , 0% | 0 , 0% | 0 , 0% | 0 , 0% |
| ε2/ε2 | 13, 0.40% | 11 , 0.38% | 0 , 0% | 0 , 0% | 0 , 0% | 0 , 0% | 0 , 0% | 1 , 0.41% | 0 , 0% | 1 , 2.50% | 0 , 0% | 0 , 0% |
| ε2/ε3 | 352, 10.76% | 310 , 10.62% | 4 , 13.79% | 1 , 7.14% | 1 , 20% | 0 , 0% | 0 , 0% | 31 , 12.60% | 0 , 0% | 4 , 10% | 0 , 0% | 1 , 25% |
| ε2/ε4 or ε1/ε3 | 49, 1.50% | 40 , 1.37% | 1 , 3.45% | 1 , 7.14% | 0 , 0% | 0 , 0% | 0 , 0% | 7 , 2.85% | 0 , 0% | 0 , 0% | 0 , 0% | 0 , 0% |
| ε3/ε3 | 2100, 64.22% | 1885 , 64.58% | 19 , 65.52% | 9 , 64.29% | 3 , 60% | 0 , 0% | 4 , 100% | 146 , 59.35% | 5 , 100% | 26 , 65% | 0 , 0% | 3 , 75% |
| ε3/ε4 | 698, 21.34% | 620 , 21.24% | 5 , 17.24% | 3 , 21.43% | 1 , 20% | 0 , 0% | 0 , 0% | 60 , 24.39% | 0 , 0% | 9 , 22.50% | 0 , 0% | 0 , 0% |
| ε4/ε4 | 54, 1.65% | 53 , 1.82% | 0 , 0% | 0 , 0% | 0 , 0% | 0 , 0% | 0 , 0% | 1 , 0.41% | 0 , 0% | 0 , 0% | 0 , 0% | 0 , 0% |
| total | 3270 | 2919 | 29 | 14 | 5 | 1 | 4 | 246 | 5 | 40 | 3 | 4 |

| 100KGP |  |  |  |  |  |  |  |
| --- | --- | --- | --- | --- | --- | --- | --- |
| Dementia |  |  |  |  |  |  |  |
| Genotypes | Number of participants (n, %) | EUR (n, %) | AFR (n, %) | AMR (n, %) | EAS (n, %) | SAS (n, %) | NA (n, %) |
| NA | 10, 5.55% | 0, 0% | 0, 0% | 0, 0% | 0, 0% | 0, 0% | 10, 100% |
| ε2/ε3 | 11, 6.11% | 9, 6.29% | 1, 14.28% | 0, 0% | 0, 0% | 1, 6.25% | 0, 0% |
| ε2/ε4 or ε1/ε3 | 3, 1.67% | 3, 2.10% | 0, 0% | 0, 0% | 0, 0% | 0, 0% | 0, 0% |
| ε3/ε3 | 69, 38.33% | 58, 40.56% | 1, 14.28% | 2, 66.67% | 1, 100% | 7, 43.75% | 0, 0% |
| ε3/ε4 | 66, 36.67% | 54, 37.76% | 4, 57.14% | 0, 0% | 0, 0% | 8, 50% | 0, 0% |
| ε4/ε4 | 21, 11.67% | 19, 13.29% | 1, 14.28% | 1, 33.33% | 0, 0% | 0, 0% | 0, 0% |
| Total | 180 | 143 | 7 | 3 | 1 | 16 | 10 |
| Controls |  |  |  |  |  |  |  |
| Genotypes | Number of participants (n, %) | EUR (n, %) | AFR (n, %) | AMR (n, %) | EAS (n, %) | SAS (n, %) | NA (n, %) |
| NA | 31, 0.89% | 0, 0% | 0, 0% | 0, 0% | 0, 0% | 0, 0% | 31, 100% |
| ε2/ε2 | 25, 0.71% | 22, 0.72% | 1, 1.61% | 1, 1.16% | 0, 0% | 1, 0.41% | 0, 0% |
| ε2/ε3 | 436, 12.53% | 394, 12.97% | 7, 11.29% | 9, 10.46% | 4, 18.18% | 22, 9.12% | 0, 0% |
| ε2/ε4 or ε1/ε3 | 72, 2.06% | 61, 2.00% | 6, 9.67% | 1, 1.16% | 1, 4.54% | 3, 1.24% | 0, 0% |
| ε3/ε3 | 2042, 58.69% | 1765, 58.11% | 25, 40.32% | 61, 70.93% | 14, 63.63% | 177, 73.44% | 0, 0% |
| ε3/ε4 | 802, 23.05% | 729, 24.00% | 20, 32.25% | 13, 15.11% | 3, 13.63% | 37, 15.35% | 0, 0% |
| ε4/ε4 | 71, 2.04% | 66, 2.17% | 3, 4.83% | 1, 1.16% | 0, 0% | 1, 0.41% | 0, 0% |
| Total | 3479 | 3037 | 62 | 86 | 22 | 241 | 31 |

In 100KGP, genotypes that could not be detected are shown as NA. AD, Alzheimer's disease; DLB, Dementia with Lewy bodies; 100KGP, 100,000 Genomes Project; ADSP, Alzheimer's Disease Sequencing Project; UKB, UK Biobank; AMP PD, Accelerating Medicines Partnership in Parkinson’s Disease; AoU, All of Us; EUR, European; AFR, African; AMR, American Admixed; AAC,African Admixed; AJ, Ashkenazi Jewish; CAS, Central Asian; EAS, Eastern Asian; SAS, South Asian; MDE, Middle Eastern; FIN, Finnish; CAH, Complex Admixture History.

When combining results across all datasets, the frequency of *APOE* ε4/ε4 in African and African Admixed AD patients is still higher than in Europeans, but the values are not significantly different. However, the frequency of *APOE* ε4/ε4 in control individuals of African and African Admixed ancestries was found to be substantially higher than in controls of European ancestry. Additionally, the frequency of *APOE* ε4/ε4 in Finnish individuals was found to be higher in AD cases and lower in controls compared to Europeans.

#### Disease-modifying variants in *APOE* ε4 carriers modulate Alzheimer’s and dementia risk across different ancestries

The summary of our findings for the frequencies of protective and disease-modifying variants under study, alongside *APOE* genotypes across all five datasets, is depicted in **Supplementary Tables 7–11**. The proportions of individuals carrying *APOE* ε4 homozygous or heterozygous genotypes alongside protective or disease-modifying variants, within the total population, total ε4/ε4 carriers, and total ε4 carriers across each ancestry, combined across all biobanks, in AD, related dementias, and controls are reported in **Figure 5, Supplementary Figure 4** and **Supplementary Table 12.** Summaries of our findings for all the assessed models in *APOE* ε4, ε4ε4, and ε3ε3 are shown in **Figure 6**, **Table 6** and **Supplementary Tables 13–22**.

**Figure 5.**
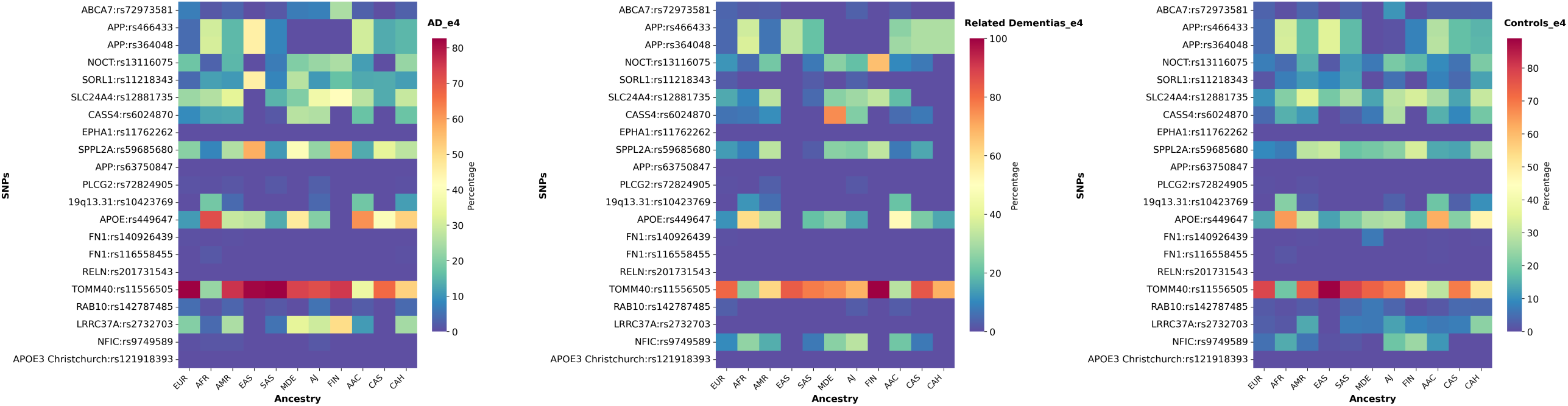
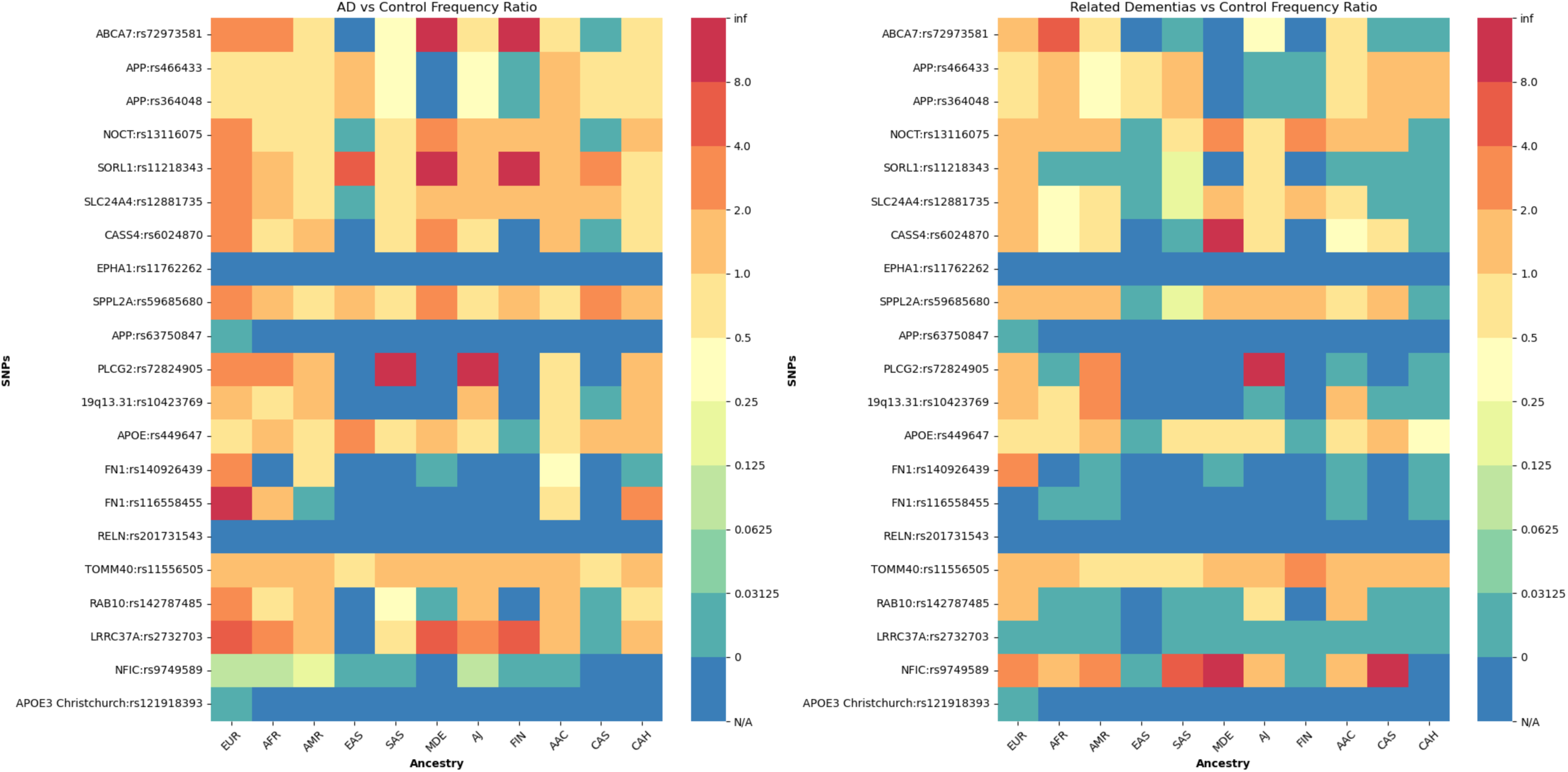
Proportions of individuals carrying both *APOE* ε4 and protective or disease- modifying variants across 11 genetic ancestries in Alzheimer’s disease, related dementias, and controls in all datasets. (A) SNP frequencies for each variant within each ancestry relative to the total number of ε4 carriers per ancestry across all datasets. (B) AD-to-control allele frequency ratios (left) and related dementia-to-control ratios (right). Warmer colors represent higher frequencies in cases versus controls, while cooler colors represent higher frequencies in controls versus cases, with dark blue (N/A) representing variants not present in either cases or controls.

**Figure 6.**
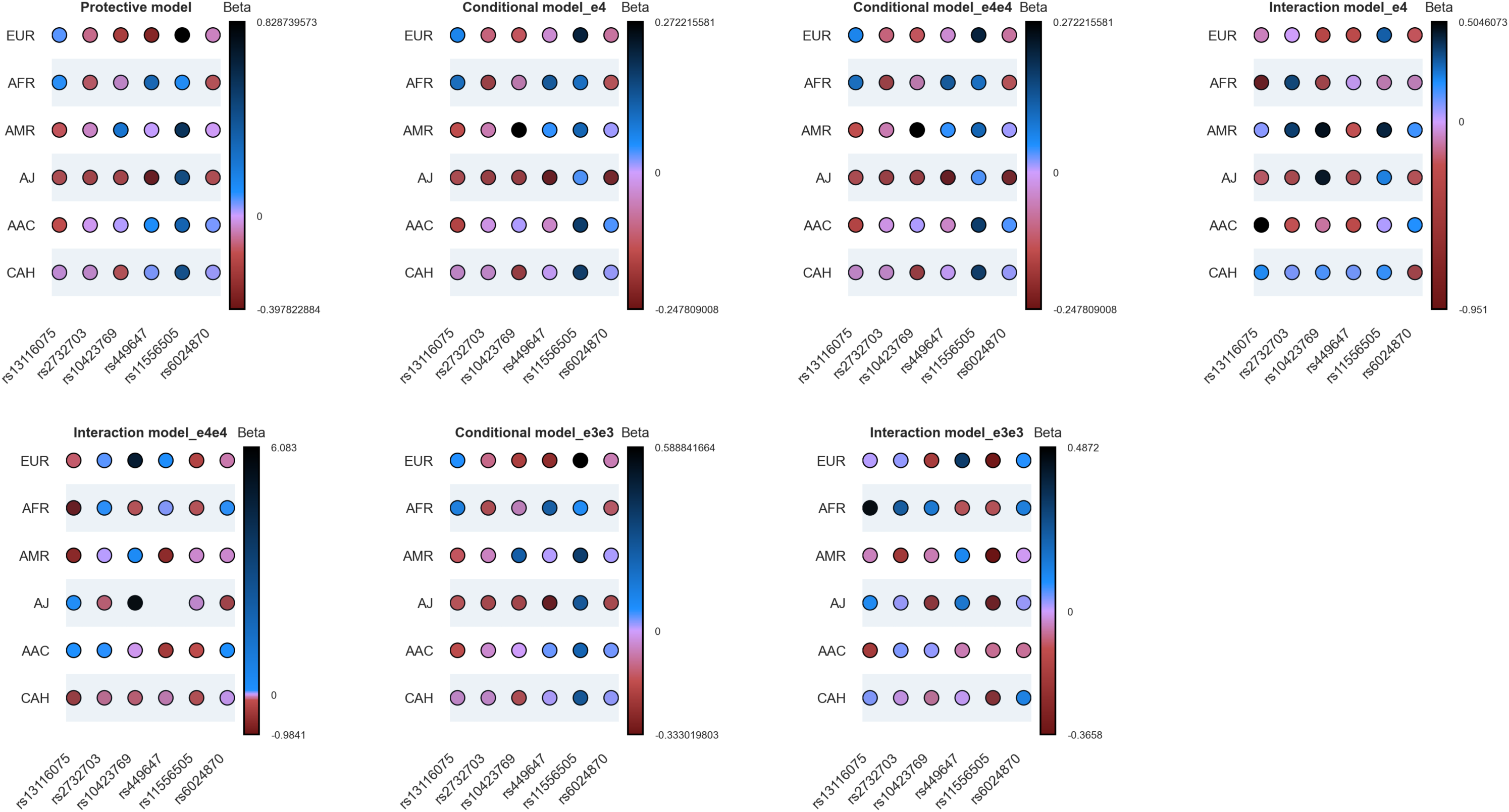
Upset plot showing protective, conditional, and interaction models across multiple ancestries. The Y-axis represents each ancestry population with a large enough sample size, and the X-axis represents the six protective/disease-modifying variants. The color bar shows the magnitude of effects as log of the odds ratio (beta value) and directionality, with red color denoting negative directionality, and blue colors denoting positive directionality.

**Table 6.**
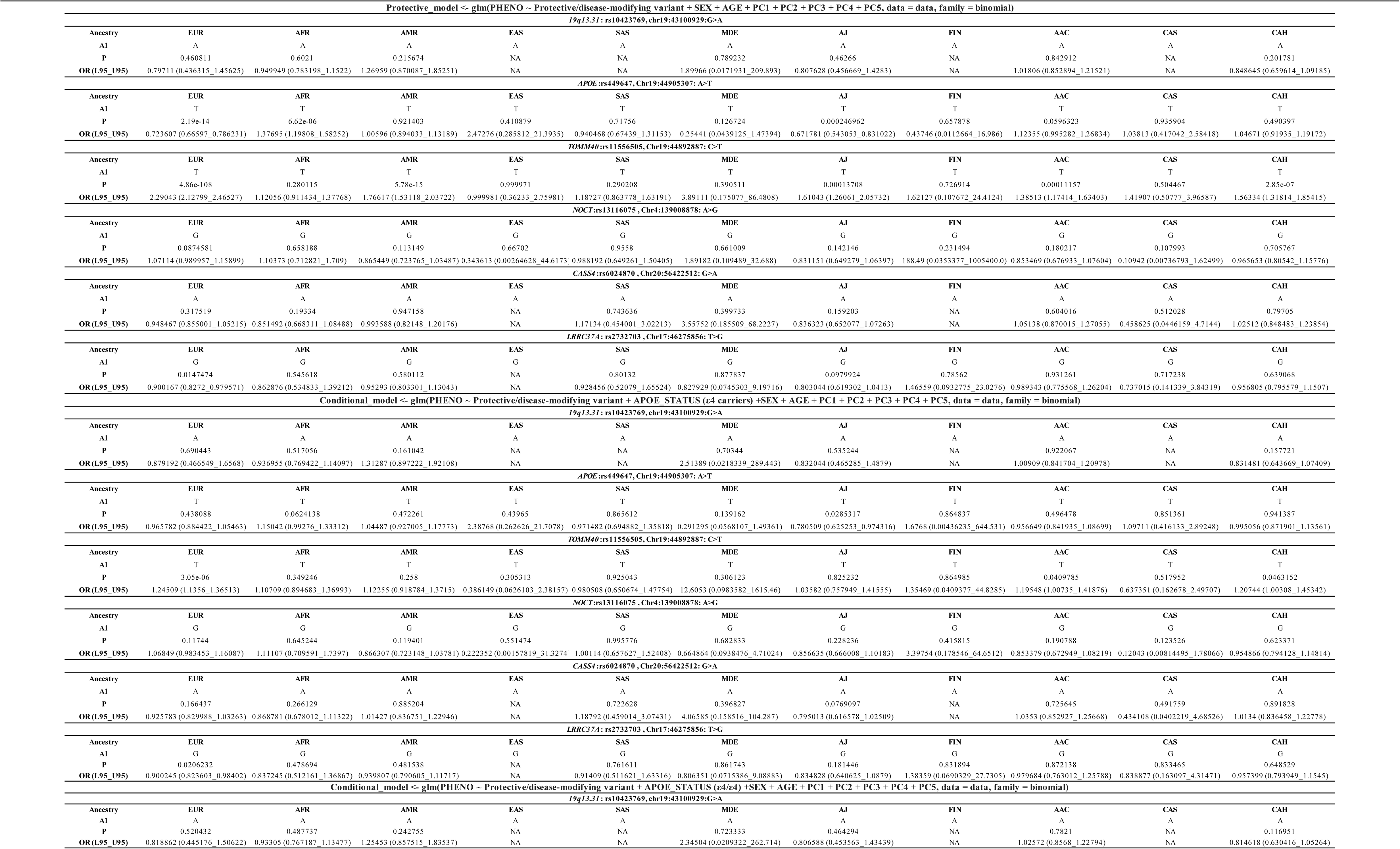
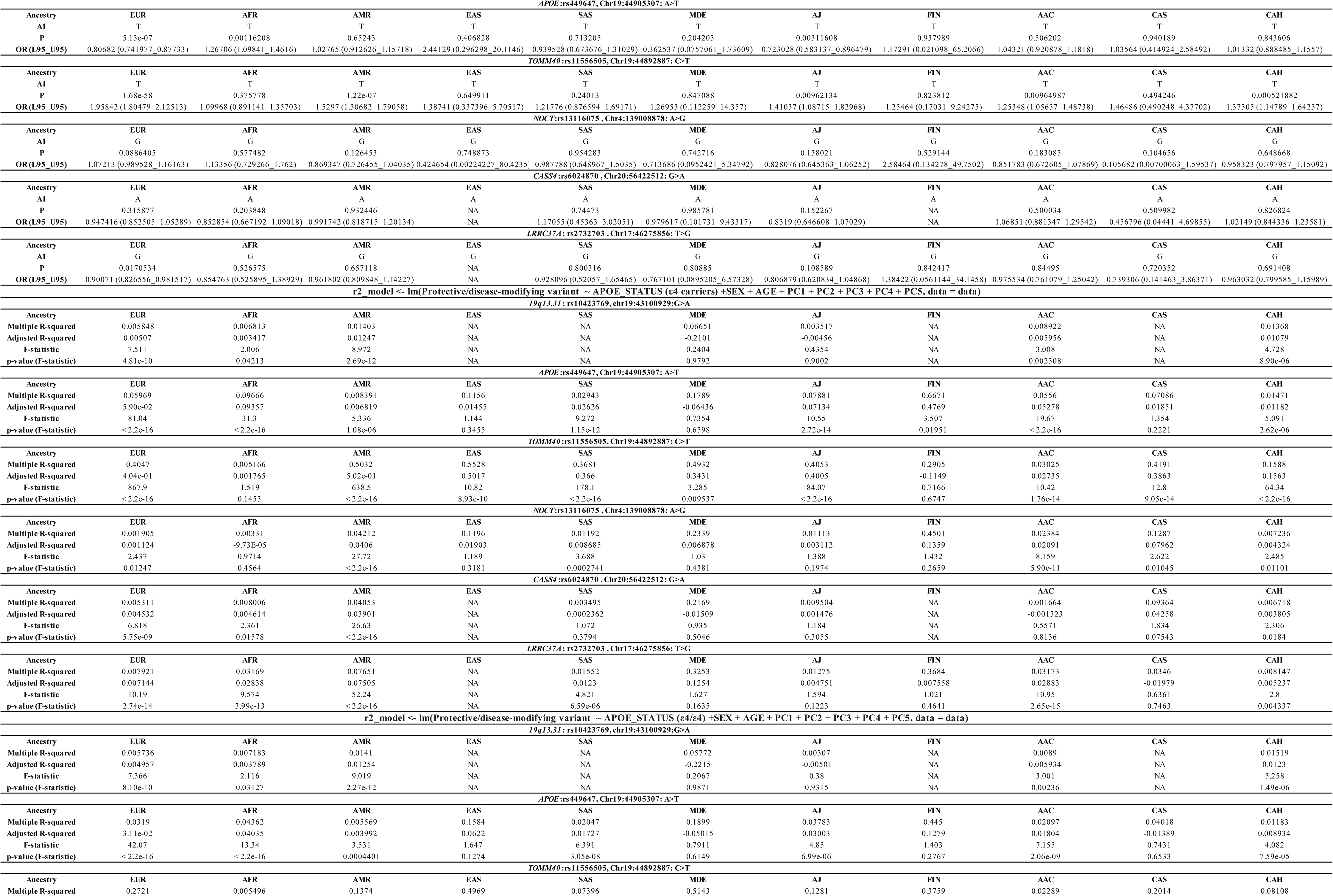
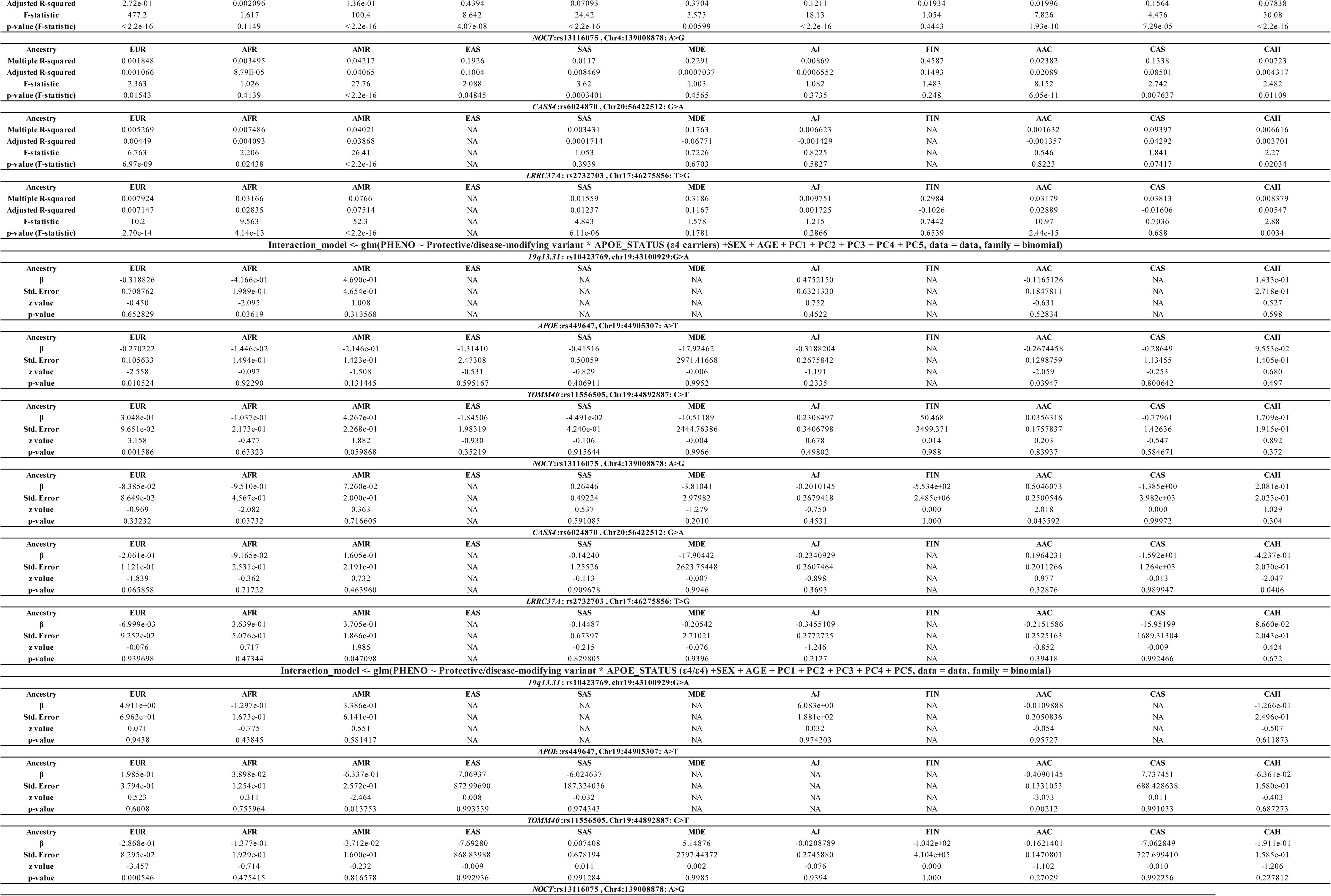
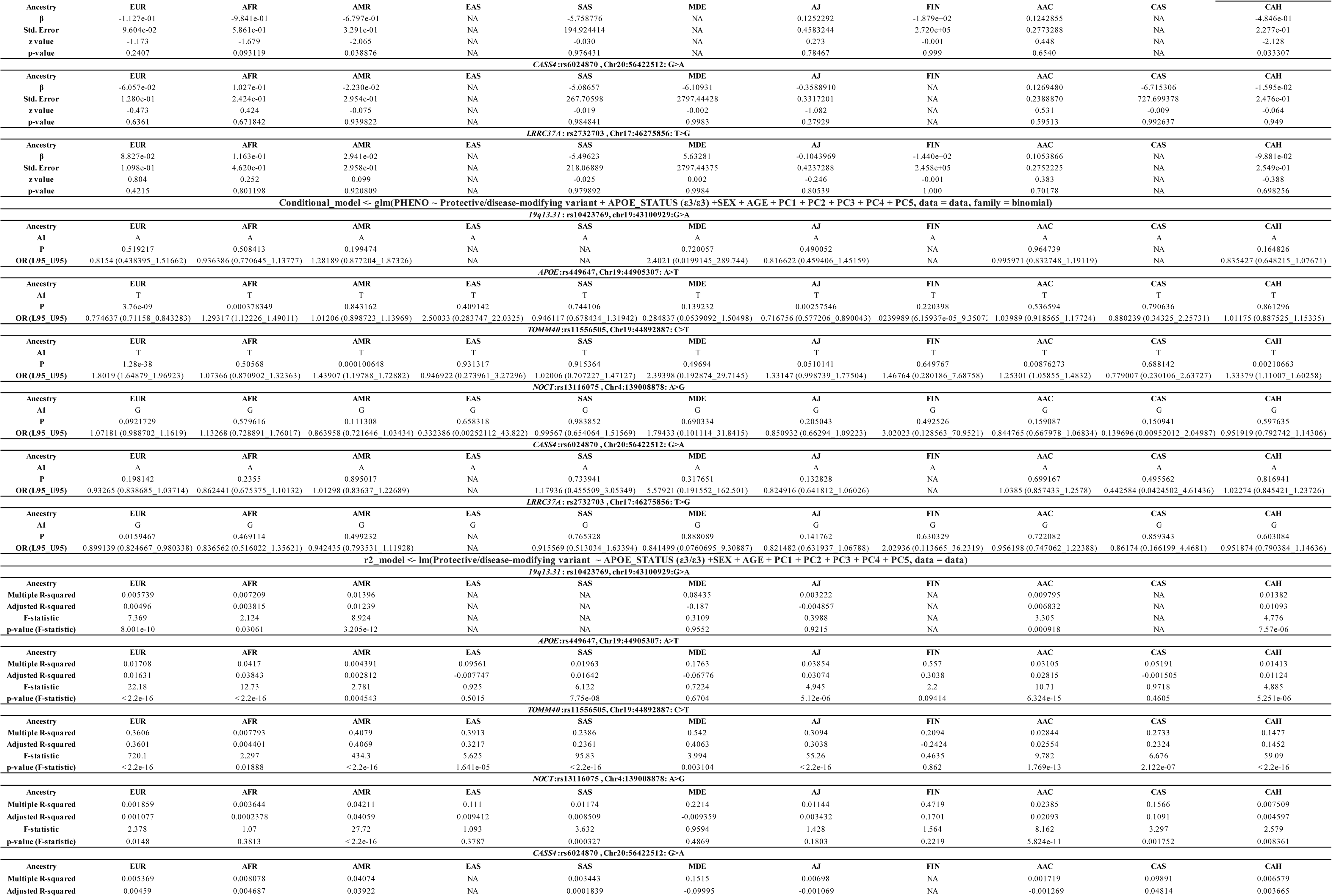
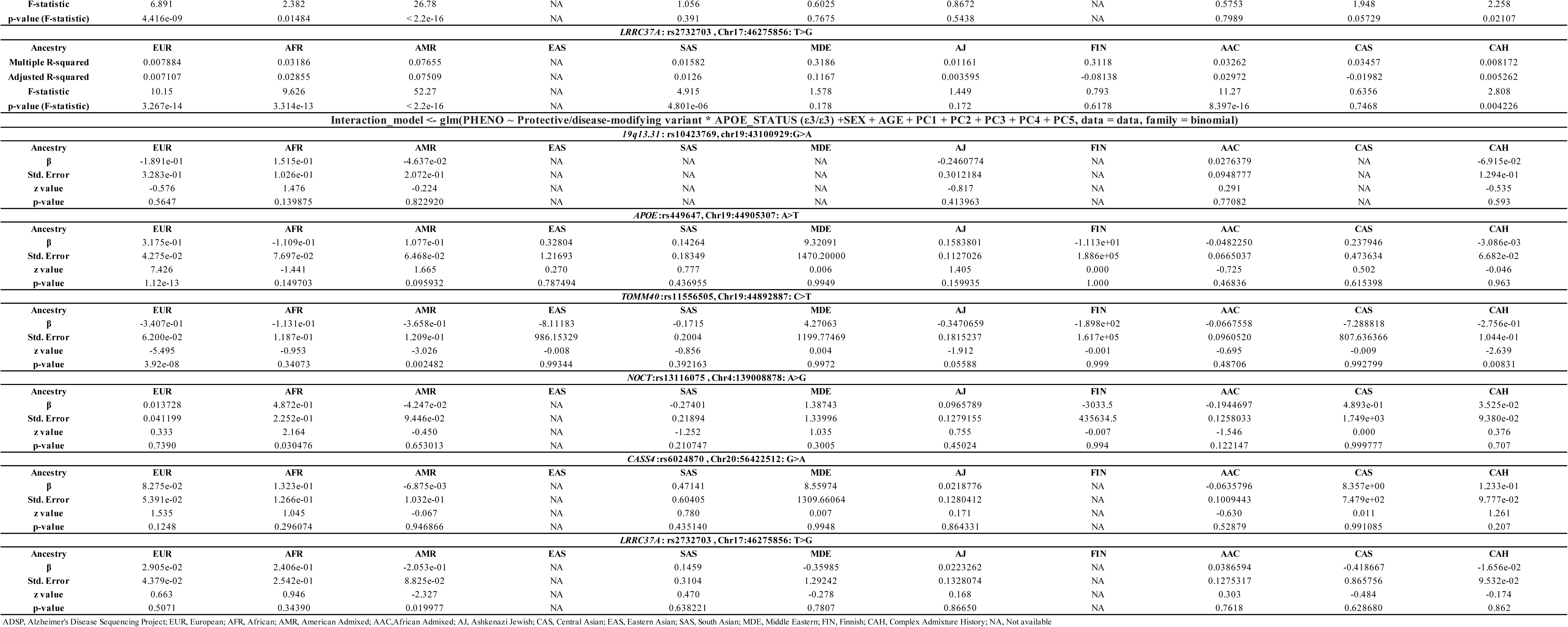
Summary of the analysis of protective/disease-modifying variants in Alzheimer’s disease cases and controls in the ADSP.

In brief, we observe higher frequencies of individuals carrying *APOE*:rs449647-T, *19q13.31*:rs10423769-A, *APP*:rs466433-G, or *APP*:rs364048-C protective variant alleles alongside either one or two copies of *APOE* ε4 among African and African Admixed ancestries compared to Europeans in AD, related dementias, and controls. Of note, carriers of *APOE*:rs449647-T and *19q13.31*:rs10423769-A are particularly noteworthy because *APOE*:rs449647-T displays the highest frequency among these ancestries, and the ratio of frequencies for *19q13.31*:rs10423769-A in both African and African Admixed ancestries compared to Europeans is substantially higher than the other protective/disease-modifying variants investigated across all three cohorts. In individuals of African ancestry, *19q13.31*:rs10423769-A was found to have a higher frequency in controls compared to both AD and related dementia cases among *APOE* ε4 homozygous or heterozygous carriers. In contrast, *APOE*:rs449647-T was found to have a lower frequency in controls compared to AD cases among *APOE* ε4 homozygous or heterozygous carriers in African ancestry but showed a higher frequency in controls carrying *APOE* ε4/ε4 versus cases in European populations (**Supplementary Table 12**).

The combination of *TOMM40*:rs11556505-T with either homozygous or heterozygous *APOE* ε4 was observed to have a higher frequency in Europeans and a lower frequency in Africans compared to most other ancestries across all three phenotypes. Additionally, the combination of *NOCT*:rs13116075-G and *APOE* ε4 homozygous or heterozygous was found to have a higher frequency in individuals of European and African Admixed ancestry versus Africans, in AD cases as compared to controls.

The protective model shows a modifying effect of *APOE*:rs449647-T in European, African, and Ashkenazi Jewish ancestries as well as a modifying effect of *TOMM40*:rs11556505-T in European, American Admixed, Ashkenazi Jewish, African Admixed, and individuals of Complex Admixture History. The R² model indicates that these variants are not in linkage disequilibrium with the *APOE* risk variants rs429358 and rs7412 **(Table 6)**.

Significant interactions were found between *APOE* ε4 and the following variants: *19q13.31*:rs10423769-A in Africans; *NOCT*:rs13116075-G in both African and African Admixed populations; *CASS4*:rs6024870-A in the Complex Admixed History group; *LRRC37A*:rs2732703- G in the American Admixed ancestry; *APOE*:rs449647-T in European and African Admixed ancestries; and *TOMM40*:rs11556505-T in Europeans. In *APOE* ε4/ε4 carriers, the interaction with *APOE*:rs449647-T was found to be significant in American Admixed and African Admixed populations, while *TOMM40*:rs11556505-T was significant in the European ancestry **(Table 6)**.

The interaction model in *APOE* ε3/ε3 shows no significant p-value for *19q13.31*:rs10423769-A in Africans, *NOCT*:rs13116075-G in African Admixed ancestry, *CASS4*:rs6024870-A in Complex Admixed History, and *LRRC37A*:rs2732703-G in American Admixed ancestry, but highly significant p-values for *APOE*:rs449647-T and *TOMM40*:rs11556505-T in European and *NOCT*:rs13116075-G in Africans with opposite directional effects compared to *APOE* ε4 carriers. These data confirm the role of these variants in modulating the effect of *APOE* ε4 in AD risk **(Table 6)**.

## Discussion

We undertook the largest and most comprehensive characterization of potential disease-causing, risk, protective, and disease-modifying variants in known AD/ADRDs genes to date, aiming to create an accessible genetic catalog of both known and novel coding and splicing variants associated with AD/ADRDs in a global context. Our results expand our understanding of the genetic basis of these conditions, potentially leading to new insights into their pathogenesis, risk, and progression. A comprehensive genetic catalog can inform the development of targeted therapies and personalized medicine approaches in the new era of precision therapeutics.

We present a user-friendly platform for the AD/ADRDs research community that enables easy and interactive access to these results (https://niacard.shinyapps.io/MAMBARD_browser/). This browser may serve as a valuable resource for researchers, clinicians, and clinical trial design.

We identified 116 genetic variants (18 known and 98 novel) in AD/ADRDs across diverse populations. The successful replication of novel variants across different datasets increases the likelihood of these variants being pathogenic and warrants further validation through future functional studies, highlighting their potential broader applicability and significance in global genetics research.

We identified 20 potentially disease-causing variants in non-European ancestries, including 13 that were absent in individuals of European ancestry. This highlights the necessity of expanding genetics research to diverse populations, corroborating the notion that the genetic architecture of AD/ADRDs risk differs across populations. We identified a total of 21 variants in control individuals that had been previously reported as disease-causing. This scenario involves three possibilities: (i) the mutation is a non-disease-causing variant found by chance in an AD patient, (ii) the mutation is pathogenic but exhibits incomplete penetrance, or (iii) control individuals represent undiagnosed patients. These findings reveal the potential for conflicting reports and misinterpretations, emphasizing the need for careful analysis and functional validation in genetics research. It underscores the critical importance of caution in identifying and interpreting potentially-pathogenic variants, which is essential for ensuring accurate diagnosis, risk assessment, genetic counseling, and development of effective treatments.

We conducted genotype-phenotype correlations for both known and novel variants. Our findings involving known variants largely reinforce previous studies, while expanding the clinical spectrum for various types of dementia, exhibiting different AAO and/or additional clinical features not previously reported. While the genotype-phenotype correlations for newly identified variants require further investigation to fully understand their impact, we identified 19 novel variants that may potentially be associated with early-onset dementia and therefore warrant further study.

While several studies have conducted *APOE* genotyping across different age groups, sexes, and population ancestries [78,79], the differential role of *APOE* across 11 ancestries in a global context has not yet been explored. We found that individuals of African and African Admixed ancestries harbor a higher frequency of *APOE* ε4/ε4 carriers than individuals of European ancestry. Recent studies have shed light on the varying risk associated with *APOE ε4* alleles in populations of African ancestry. Indeed, it has been reported that individuals of African descent who carry the *APOE ε4* allele have a lower risk of developing AD compared to other populations with the same allele. This suggests that the genetic background of African ancestry around the *APOE* gene is linked to a reduced odds ratio for risk variants [77]. Furthermore, a recent study has suggested the presence of a resilient locus (*19q13.31)* potentially modifying *APOE ε4* risk in African-descent populations. This disease-modifying locus, located 2MB from *APOE*, significantly lowers the AD risk for African *APOE ε4* carriers, reducing the magnitude of the effect from 7.2 to 2.1 [10]. Our finding is in concordance with the largest AD meta-analysis conducted to date [77]. We identified several variants with high frequency among *APOE* ε4 homozygous or heterozygous carriers in African and African Admixed ancestries. Notably, individuals carrying both *APOE* ε4 homozygous or heterozygous and either *APOE*:rs449647-T or *19q13.31*:rs10423769-A exhibit higher frequencies in African and African Admixed ancestries compared to Europeans.

Considering all the models under study, we find that in the presence of *APOE* ε4, *APOE*:rs449647- T decreases the risk of AD in Europeans but increases it in Africans. *TOMM40*:rs11556505-T increases the risk of AD in Europeans. *TOMM40*:rs11556505-T also increases the risk of AD in American Admixed and Ashkenazi Jewish ancestries, though not through an interaction with *APOE*. An interaction effect with *APOE* was found for *19q13.31*:rs10423769-A, *NOCT*:rs13116075-G, *CASS4*:rs6024870-A, and *LRRC37A*:rs2732703-G. Specifically, *19q13.31*:rs10423769-A reduces the risk of AD in Africans, *NOCT*:rs13116075-G reduces the risk in Africans but increases it in African Admixed ancestry, *CASS4*:rs6024870-A reduces the risk in Complex Admixed History ancestry, and *LRRC37A*:rs2732703-G increases the risk in American Admixed ancestry.

Our findings support previous literature, which indicates that *19q13.31* is an African-ancestry-specific locus that reduces the risk effect of *APOE* ε4 for developing AD. *APOE*:rs449647-T is a polymorphism in the regulatory region of *APOE* that can modulate the risk of developing AD by altering its affinity to transcription factors, thus affecting gene expression. Our study demonstrates its association with an increased risk of AD in *APOE* ε4 carriers of African ancestry and a decreased risk in *APOE* ε4 carriers of European ancestry. *TOMM40*:rs11556505-T has been variably associated with both risk and protective effects, likely depending on the phenotype being evaluated [11]. We show an association with an increased risk of AD in *APOE* ε4 carriers, particularly in Europeans. In addition, this study reveals the disease-modifying effect of *NOCT*:rs13116075-G, *CASS4*:rs6024870-A and *LRRC37A*:rs2732703-G across different ancestries. The interaction of these variants with *APOE* ε4 is not known, but identifying the mechanism(s) conferring protection could provide greater insights into the etiology of AD and inform potential ancestry-specific therapeutic interventions.

This comprehensive genetic characterization, the largest of its kind for AD/ADRDs across diverse populations, holds critical implications for potential clinical trials and therapeutic interventions in a global context, highlighting its significance for such efforts worldwide. For example, clinical trials for *GRN* have recently commenced [80] (https://www.theaftd.org/posts/1ftd-in-the-news/b-ftd-grn-gene-therapy-abio/). Understanding population-specific frequencies of genetic contributors to disease is vital in the design and implementation of clinical trials for several reasons. Firstly, it allows for targeted recruitment, ensuring that studies include an adequate number of participants concordant with their genetic makeup. Secondly, it promotes diversity in clinical trial populations, which is essential to understand disease globally, as well as treatment responses across different groups. Furthermore, knowledge of population-specific variant frequencies may inform treatment efficacy assessments in the future, as genetics y influence treatment outcomes. In summary, it plays a vital role in personalized medicine, guiding more targeted and effective treatments based on individuals’ genetic profiles.

Despite our efforts, there are several limitations and shortcomings to consider in this study. A major limitation is the underrepresentation of certain populations, which leads to underpowered datasets limiting the possibility of drawing firm conclusions. Additionally, the reliance on currently available datasets may introduce biases, as these datasets often have varying levels of coverage, quality, and accurate clinical information. Another relevant consideration is the differing exclusion criteria for controls across cohorts, which may affect the comparability of results. Future research should aim to include more diverse populations and improve the quality of genetic data in addition to standardized data harmonization efforts. Moreover, functional validation of identified variants is necessary to confirm their pathogenicity and relevance to AD/ADRDs.

Lastly, ongoing collaborations between researchers, clinicians, and policy-makers are crucial to ensure that advancements in genetic research translate into equitable and effective clinical applications. Our study is a step towards addressing these limitations by providing a more diverse genetic characterization and highlighting the need for inclusive research practices. Future directions should continue to focus on enhancing the robustness and applicability of genetic findings in AD/ADRDs research, making knowledge globally relevant.

## Acknowledgments

We thank Paige Brown Jarreau for her meticulous editing of this manuscript.

This work was supported in part by the Intramural Research Program of the NIH, the National Institute on Aging (NIA), National Institutes of Health, Department of Health and Human Services; project number ZO1 AG000535 and ZIA AG000949. This work utilized the computational resources of the NIH HPC Biowulf cluster. (http://hpc.nih.gov).

The All of Us Research Program is supported by the National Institutes of Health, Office of the Director: Regional Medical Centers: 1 OT2 OD026549; 1 OT2 OD026554; 1 OT2 OD026557; 1 OT2 OD026556; 1 OT2 OD026550; 1 OT2 OD 026552; 1 OT2 OD026553; 1 OT2 OD026548; 1 OT2 OD026551; 1 OT2 OD026555; IAA #: AOD 16037; Federally Qualified Health Centers: HHSN 263201600085U; Data and Research Center: 5 U2C OD023196; Biobank: 1 U24 OD023121; The Participant Center: U24 OD023176; Participant Technology Systems Center: 1 U24 OD023163; Communications and Engagement: 3 OT2 OD023205; 3 OT2 OD023206; and Community Partners: 1 OT2 OD025277; 3 OT2 OD025315; 1 OT2 OD025337; 1 OT2 OD025276. In addition, the All of Us Research Program would not be possible without the partnership of its participants.

This research has been conducted using the UK Biobank Resource under application number 33601.

This research was made possible through access to data in the National Genomic Research Library, which is managed by Genomics England Limited (a wholly owned company of the Department of Health and Social Care). The National Genomic Research Library holds data provided by patients and collected by the NHS as part of their care and data collected as part of their participation in research. The National Genomic Research Library is funded by the National Institute for Health Research and NHS England. The Wellcome Trust, Cancer Research UK and the Medical Research Council have also funded research infrastructure.

This research has been conducted using the Alzheimer’s Disease Sequencing Project (ADSP) Resource under accession number NG00067. The data for this study were prepared, archived, and distributed by the National Institute on Aging Alzheimer’s Disease Data Storage Site (NIAGADS) at the University of Pennsylvania (U24-AG041689), funded by the National Institute on Aging.

This work was supported in part by the Intramural Research Program of the National Institute on Aging and the National Institute of Neurological Disorders and Stroke (project number Z01- AG000949-02). Data used in the preparation of this article were obtained from the AMP PD Knowledge Platform. For up-to-date information on the study, please visit https://www.amp-pd.org. AMP PD—a public-private partnership—is managed by the FNIH and funded by Celgene, GSK, the Michael J. Fox Foundation for Parkinson’s Research, the National Institute of Neurological Disorders and Stroke, Pfizer, Sanofi, and Verily. Clinical data and biosamples used in the preparation of this article were obtained from the Parkinson’s Progression Markers Initiative (PPMI), and the Parkinson’s Disease Biomarkers Program (PDBP). PPMI—a public-private partnership—is funded by the Michael J. Fox Foundation for Parkinson’s Research and funding partners, including full names of all of the PPMI funding partners found at http://www.ppmi-info.org/fundingpartners. The PPMI Investigators have not participated in reviewing the data analysis or content of the manuscript. For up-to-date information on the study, visit http://www.ppmi-info.org. The Parkinson’s Disease Biomarker Program (PDBP) consortium is supported by the National Institute of Neurological Disorders and Stroke (NINDS) at the National Institutes of Health. A full list of PDBP investigators can be found at https://pdbp.ninds.nih.gov/policy. The PDBP Investigators have not participated in reviewing the data analysis or content of the manuscript. PDBP sample and clinical data collection is supported under grants by NINDS: U01NS082134, U01NS082157, U01NS082151, U01NS082137, U01NS082148, and U01NS082133.

## Author Contributions

JDR, CB, MAN, AS and SBC, contributed to the study concept or design. MK, FA, SMG, SCA, PSL, FF, HL, JJK, MJK and MBM, were involved in the analysis of data across different biobanks. All authors contributed to the critical review and had final responsibility for the decision to submit for publication.

## Data and code availability

The code used in this study can be found online at https://github.com/NIH-CARD/ADRD-GeneticDiversity-Biobanks,10.5281/zenodo.13363465.

---

**Supplementary Figure 1- PCA plots in (A) All of Us, (B) UKB, (C) ADSP, (D) AMP PD, and (E) 100 KGP.**

**Supplementary Figure 2- Heatmaps showing the frequencies of all identified variants in the discovery and replication phases across all ancestries in each biobank.**

**Supplementary Figure 3- Proportion of *APOE* genotypes in (A) Alzheimer’s disease, (B) related dementias, and (C) controls across 11 genetic ancestries.**

Unknown genotypes and those absent across all ancestries were excluded from the analysis. Genotypes and ancestries not available in the 100KGP were also excluded.

**Supplementary Figure 4- Proportions of individuals carrying both *APOE* ε4/ε4 genotypes and protective or disease-modifying variants across 11 genetic ancestries in Alzheimer’s disease, related dementias, and controls in all datasets.**

Supplementary Figures 4A and 4C represent SNP distribution within each cohort, and Supplementary Figures 4B and 4D represent SNP distribution between cohorts. The total populations of each ancestry were used to generate 4A and 4B, while the total numbers of ε4/ε4 carriers for each ancestry were used to generate 4C and 4D. Supplementary Figures 4B and 4D show allele frequency ratios (AD-to-Control, left; Related dementias-to-Control, right) among *APOE* ε4/ε4 carriers for each of the candidate protective or disease-modifying variant, per ancestry. Warmer colors represent higher frequencies in cases versus controls, while cooler colors represent higher frequencies in controls versus cases, with dark blue (N/A) representing variants not present in either cases or controls.

Supplementary Table 1- Discovery phase: Multi-ancestry summary of all variants identified in Alzheimer’s disease and related dementia cases in AoU

Supplementary Table 2- Discovery phase: Multi-ancestry summary of all variants identified in Alzheimer’s disease and related dementia cases in UKB

Supplementary Table 3- Discovery phase: Multi-ancestry summary of all variants identified in Alzheimer’s disease and related dementia cases in 100KGP

Supplementary Table 4- Multi-ancestry summary of variants previously reported as potential disease-causing but identified in controls in multiple databases in this study

Supplementary Table 5- Phenotypic data for all individuals carrying known and novel potential disease-causing variants in the discovery phase

Supplementary Table 6- Phenotypic data for all individuals carrying known and novel potential disease-causing variants in the replication phase

Supplementary Table 7- Multi-ancestry summary of individuals carrying both *APOE* genotypes and protective or disease-modifying variants in patients and controls in AoU

Supplementary Table 8- Multi-ancestry summary of individuals carrying both *APOE* genotypes and protective or disease-modifying variants in patients and controls in ADSP

Supplementary Table 9- Multi-ancestry summary of individuals carrying both *APOE* genotypes and protective or disease-modifying variants in patients and controls in AMP PD

Supplementary Table 10- Multi-ancestry summary of individuals carrying both *APOE* genotypes and protective or disease-modifying variants in patients and controls in 100KGP

Supplementary Table 11- Multi-ancestry summary of individuals carrying both *APOE* genotypes and protective or disease-modifying variants in patients and controls in UKB

Supplementary Table 12- Combined results of data for individuals carrying both *APOE* genotypes and protective or disease-modifying variants in patients and controls across AoU, UKB, ADSP, 100KGP, and AMP PD

Supplementary Table 13- Assessment of the protective model in ADSP

Supplementary Table 14- Assessment of the conditional model for *APOE* ε4 carriers in ADSP

Supplementary Table 15- Assessment of the conditional model for *APOE* ε4/ε4 genotype in ADSP

Supplementary Table 16- Assessment of the correlation model for *APOE* ε4 carriers in ADSP

Supplementary Table 17- Assessment of the correlation model for *APOE* ε4/ε4 genotype in ADSP

Supplementary Table 18- Assessment of the interaction model for *APOE* ε4 carriers in ADSP

Supplementary Table 19- Assessment of the interaction model for *APOE* ε4/ε4 genotype in ADSP

Supplementary Table 20- Assessment of the conditional model for *APOE* ε3/ε3 genotype in ADSP

Supplementary Table 21- Assessment of the correlation model for *APOE* ε3/ε3 genotype in ADSP

Supplementary Table 22- Assessment of the interaction model for *APOE* ε3/ε3 genotype in ADSP

