## Supplementary figures and images for "Biobank-scale characterization of Alzheimer’s disease and related dementias identifies potential disease-causing variants, risk factors, and genetic modifiers across diverse ancestries"

### Supplementary Figure 1A

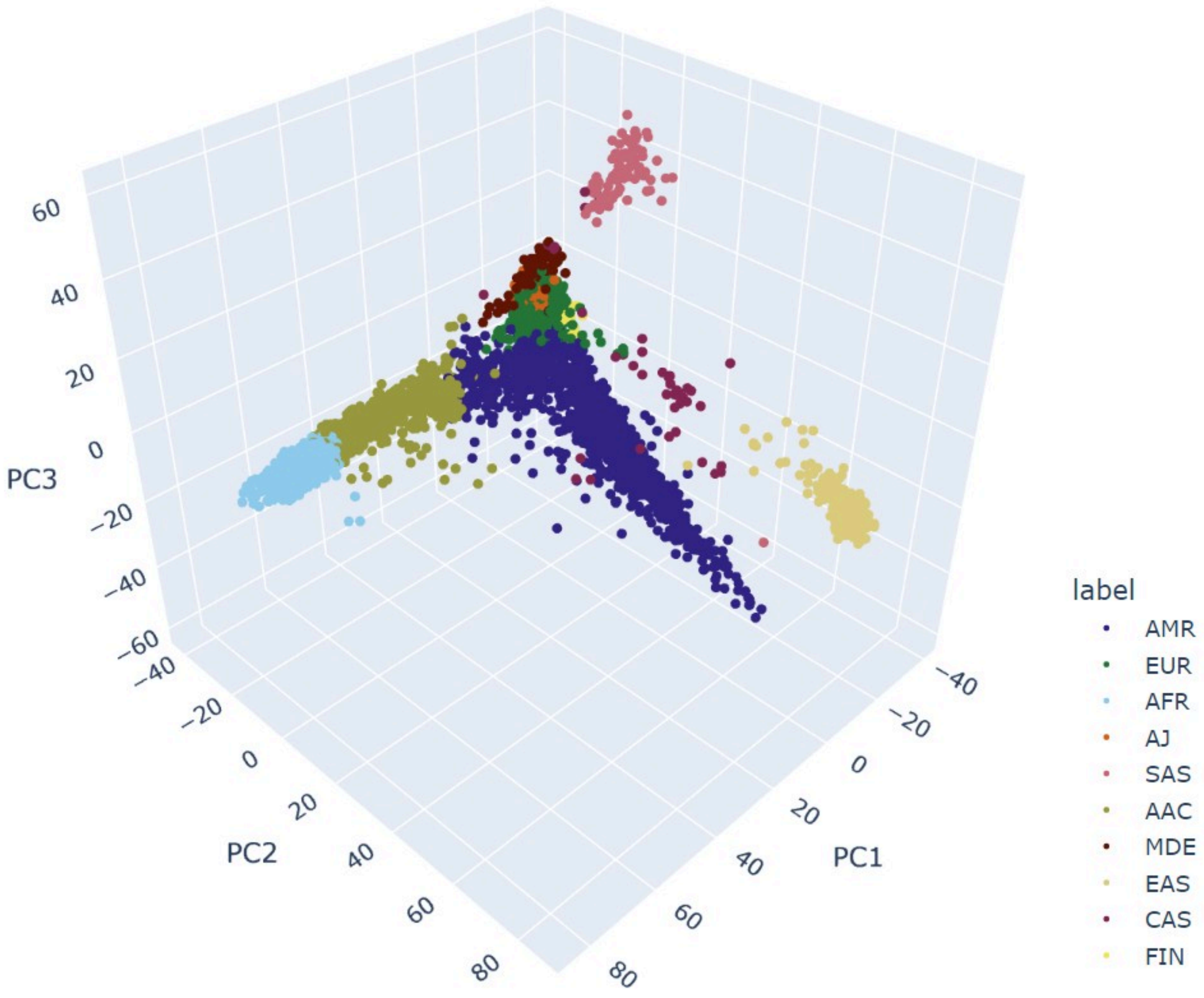

### Supplementary Figure 1B

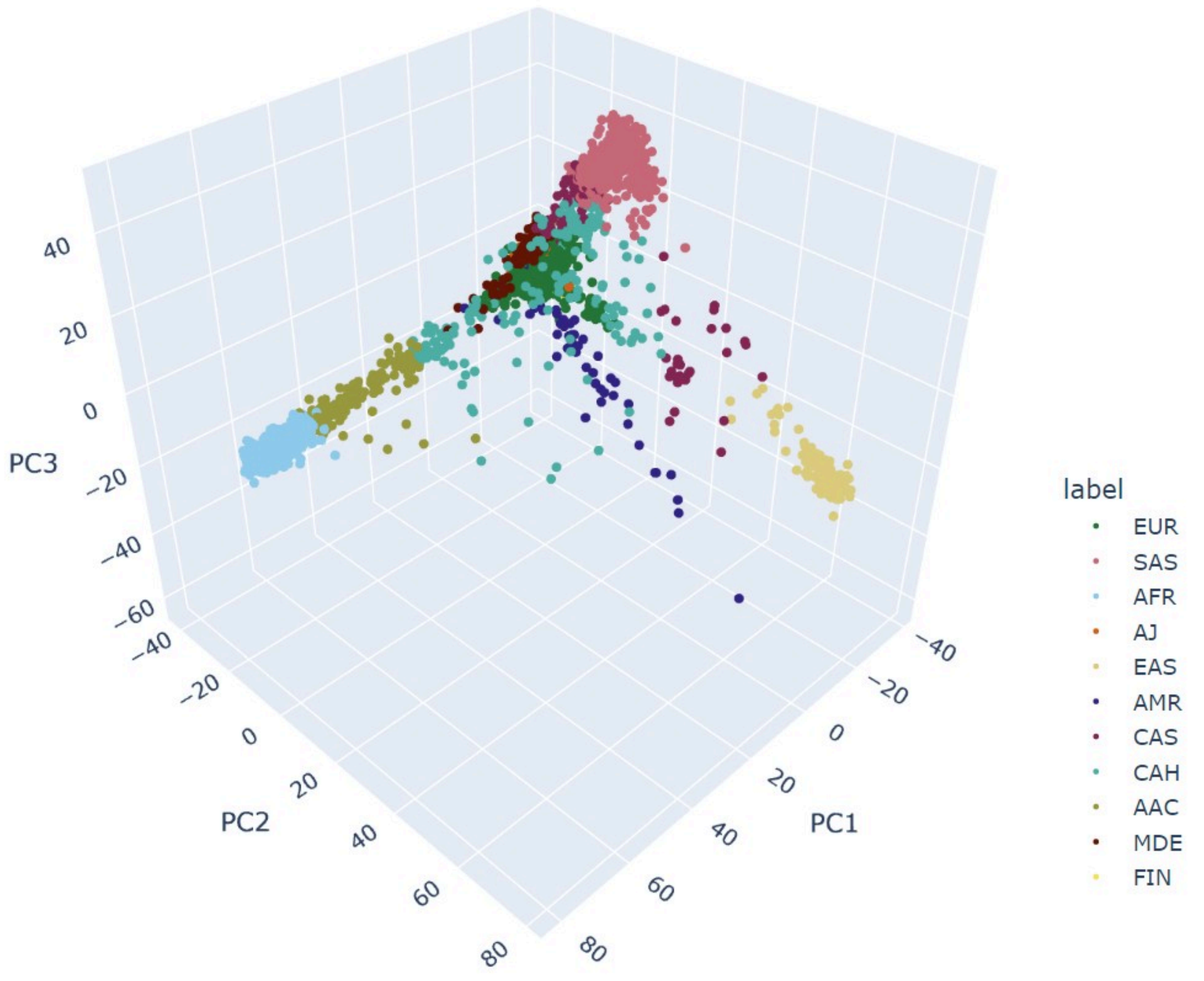

### Supplementary Figure 1C

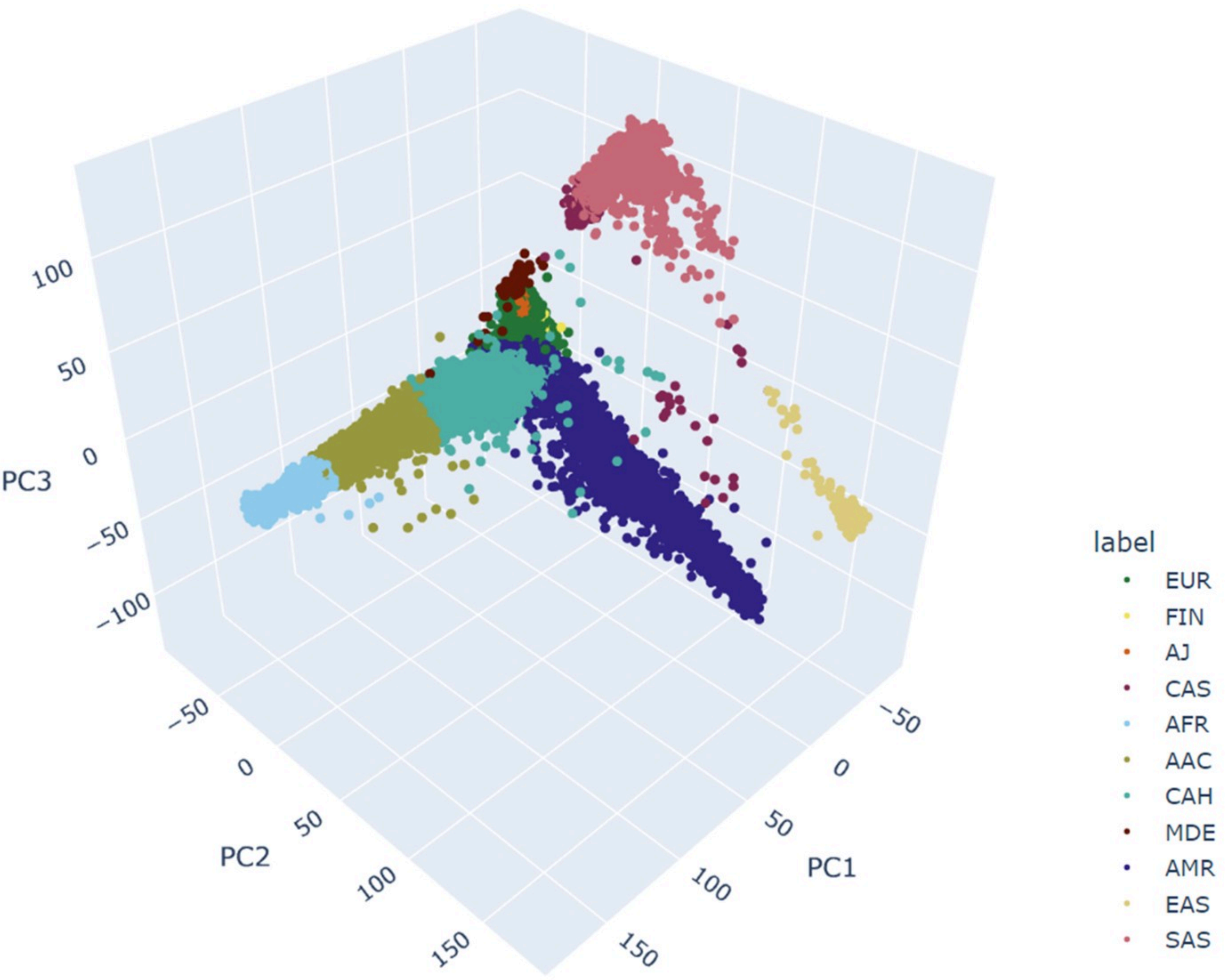

### Supplementary Figure 1D

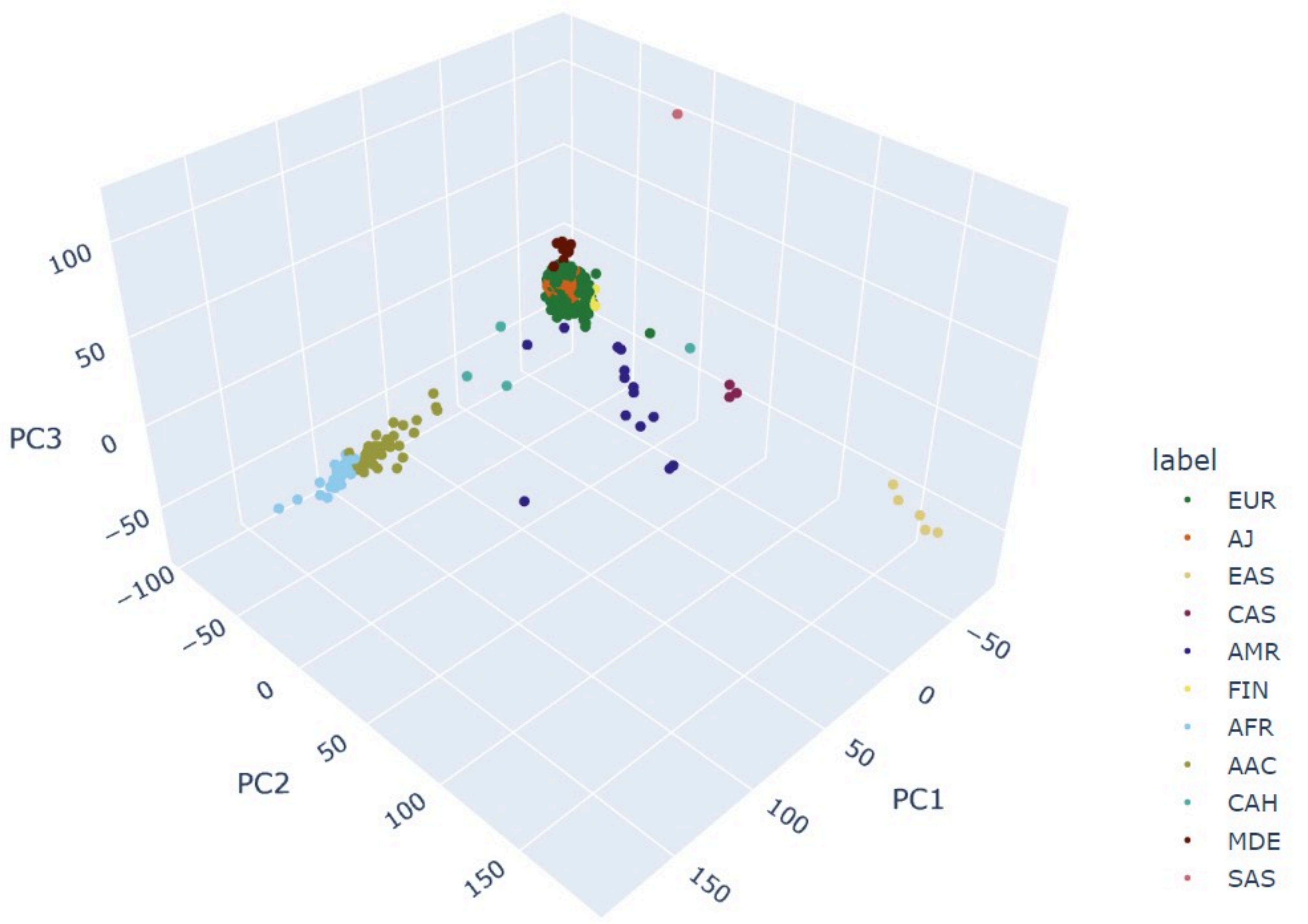

### Supplementary Figure 1E

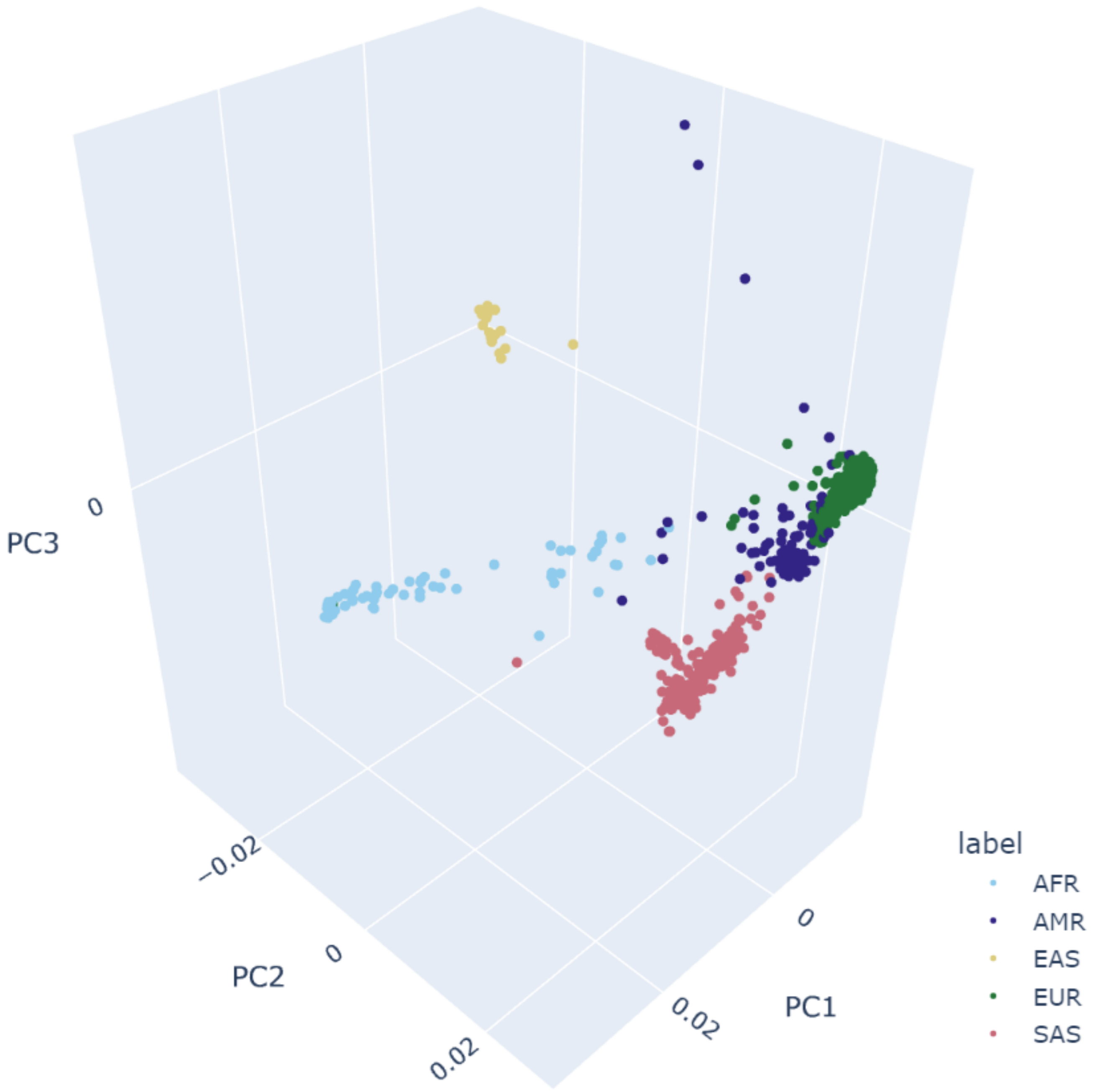

### Supplementary Figure 2

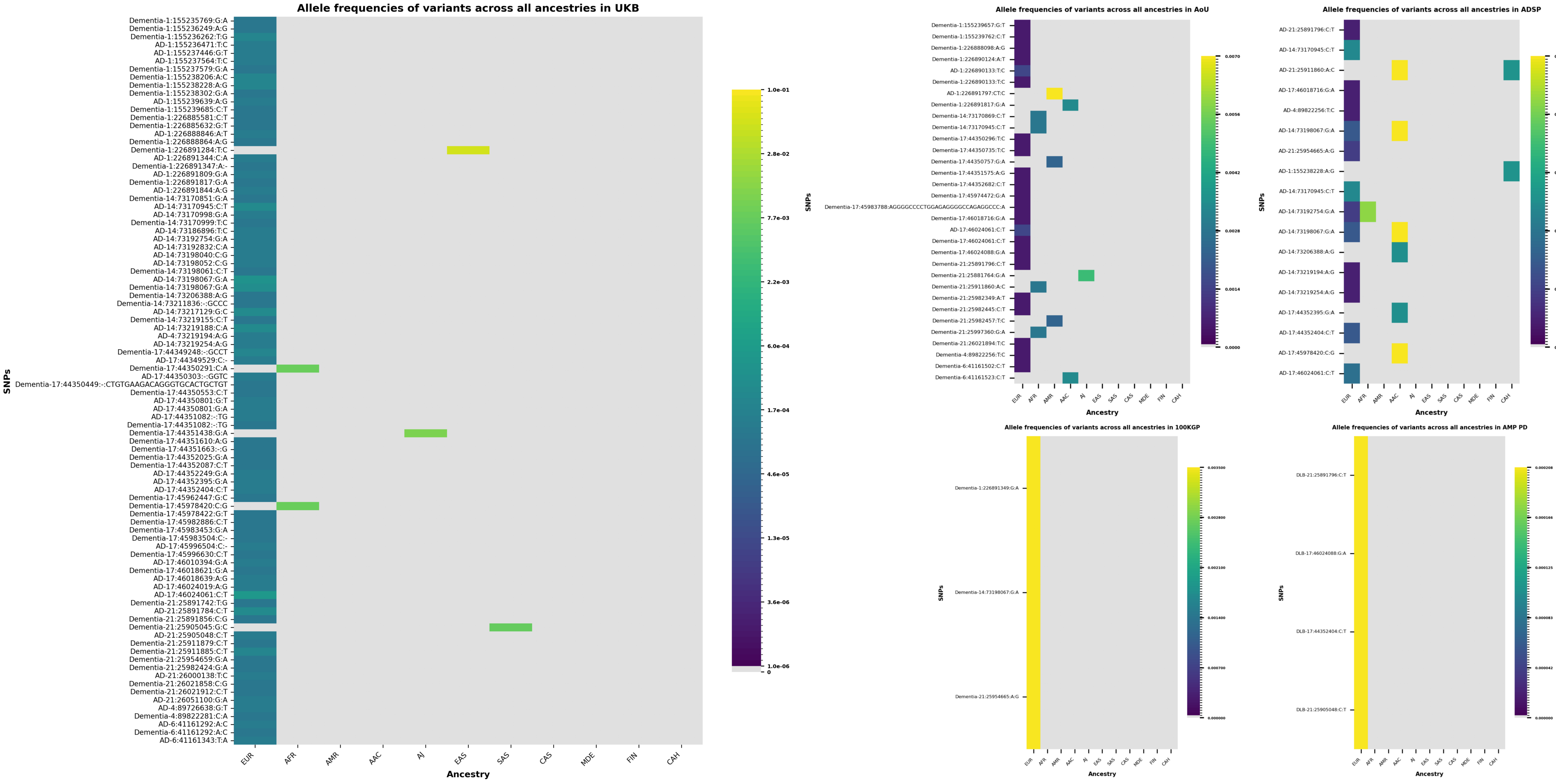

### Supplementary Figure 3A

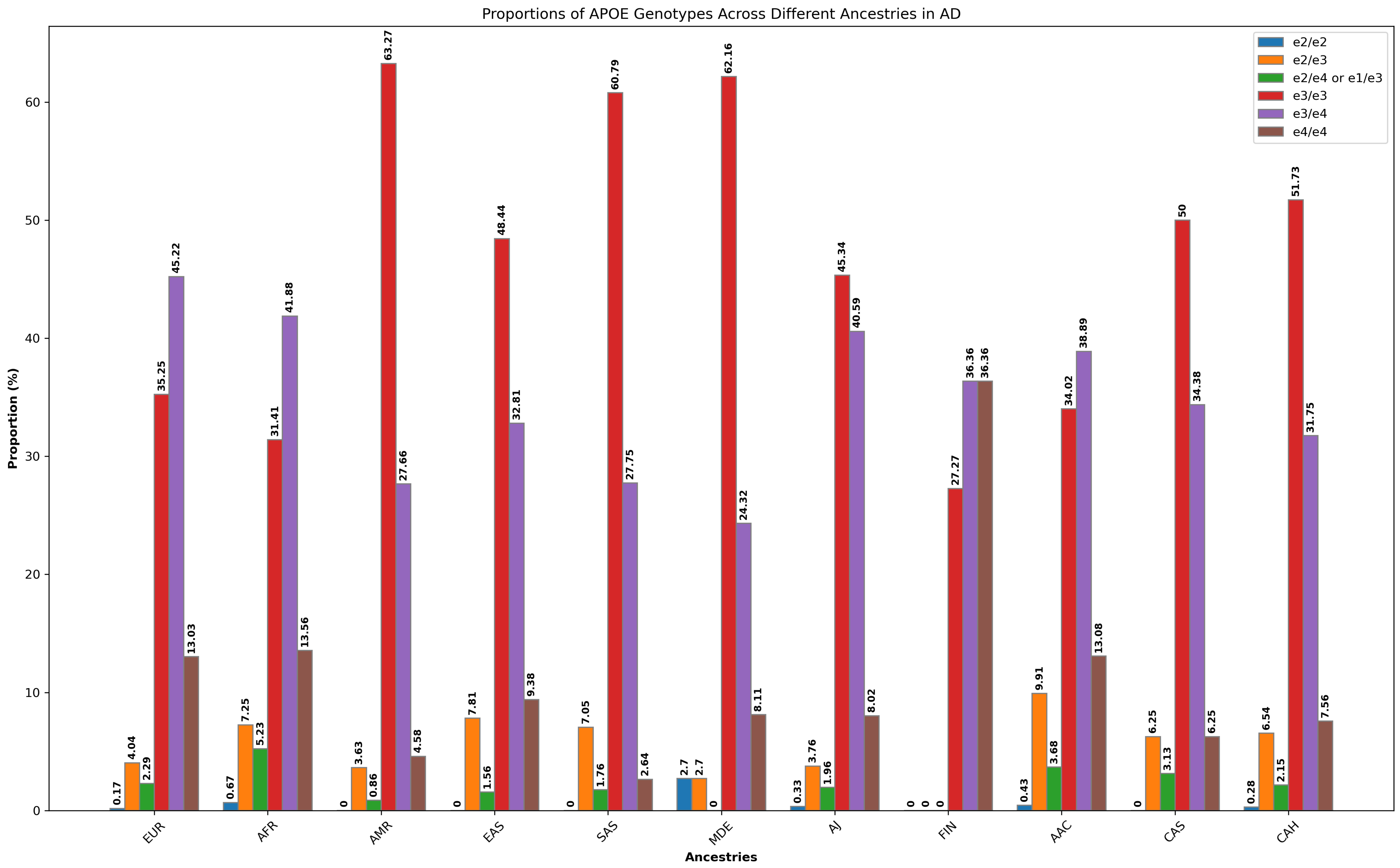

### Supplementary Figure 3B

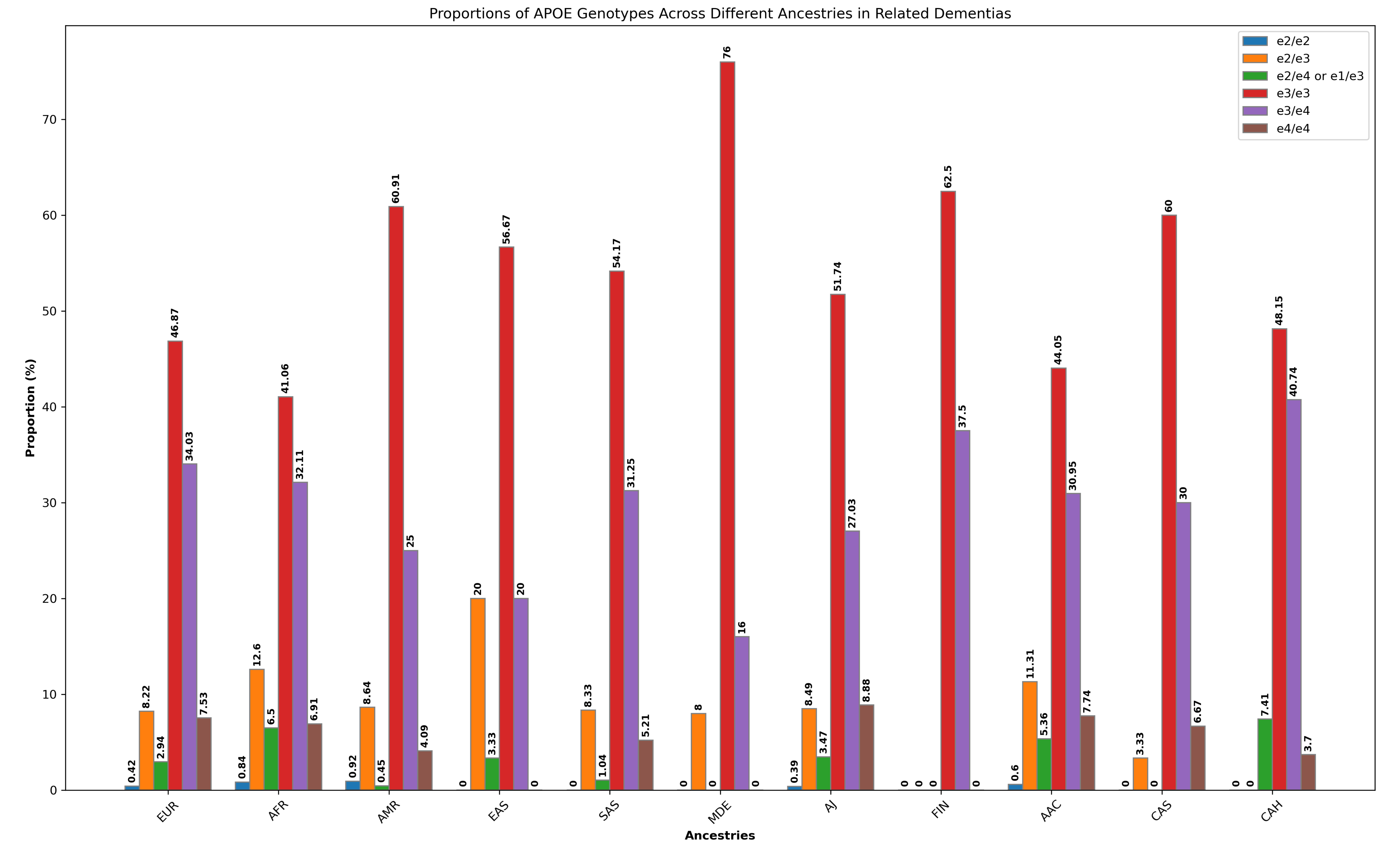

### Supplementary Figure 3C

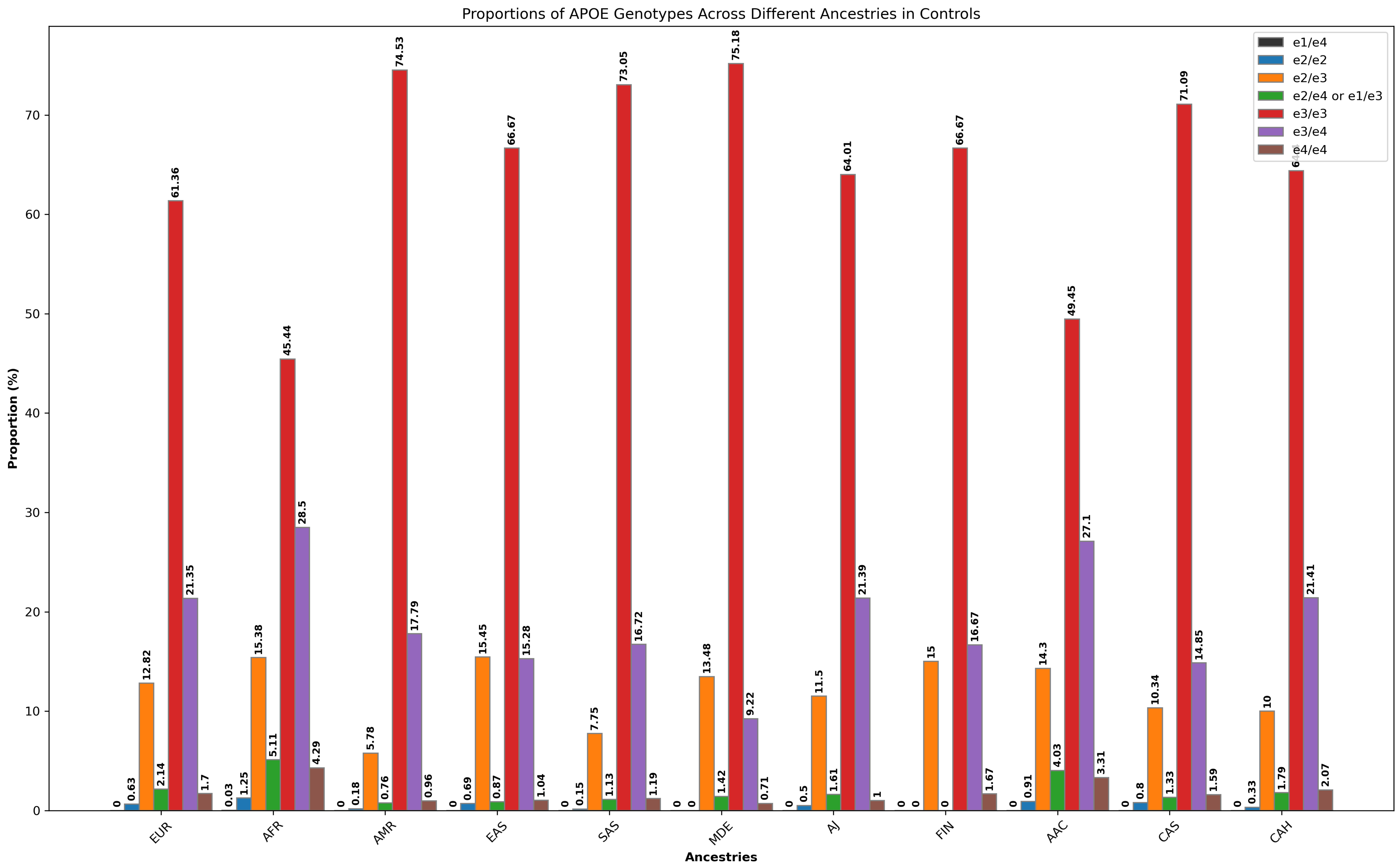

### Supplementary Figure 4A

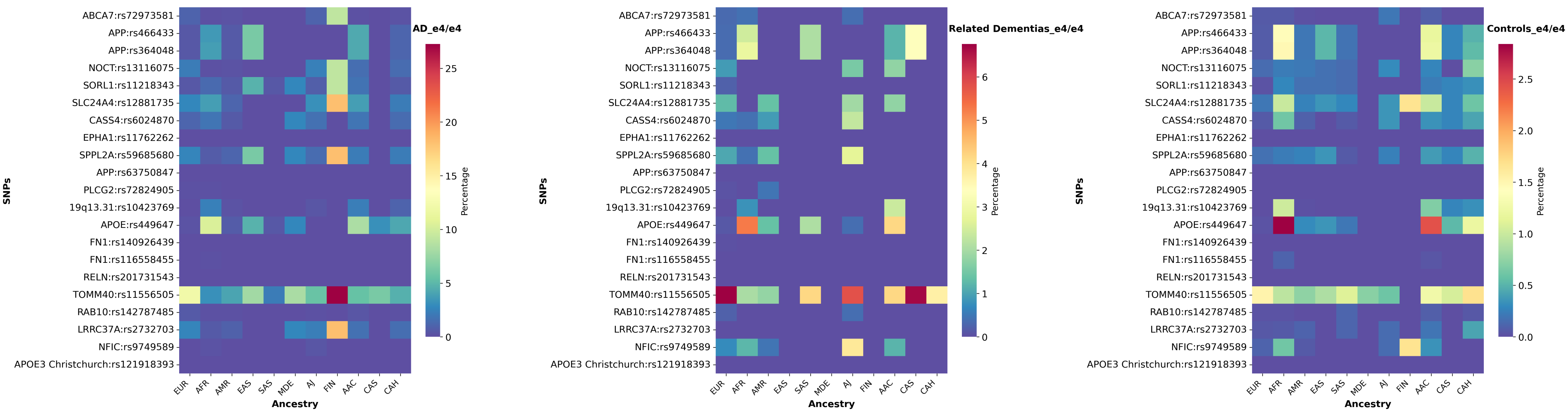

### Supplementary Figure 4B

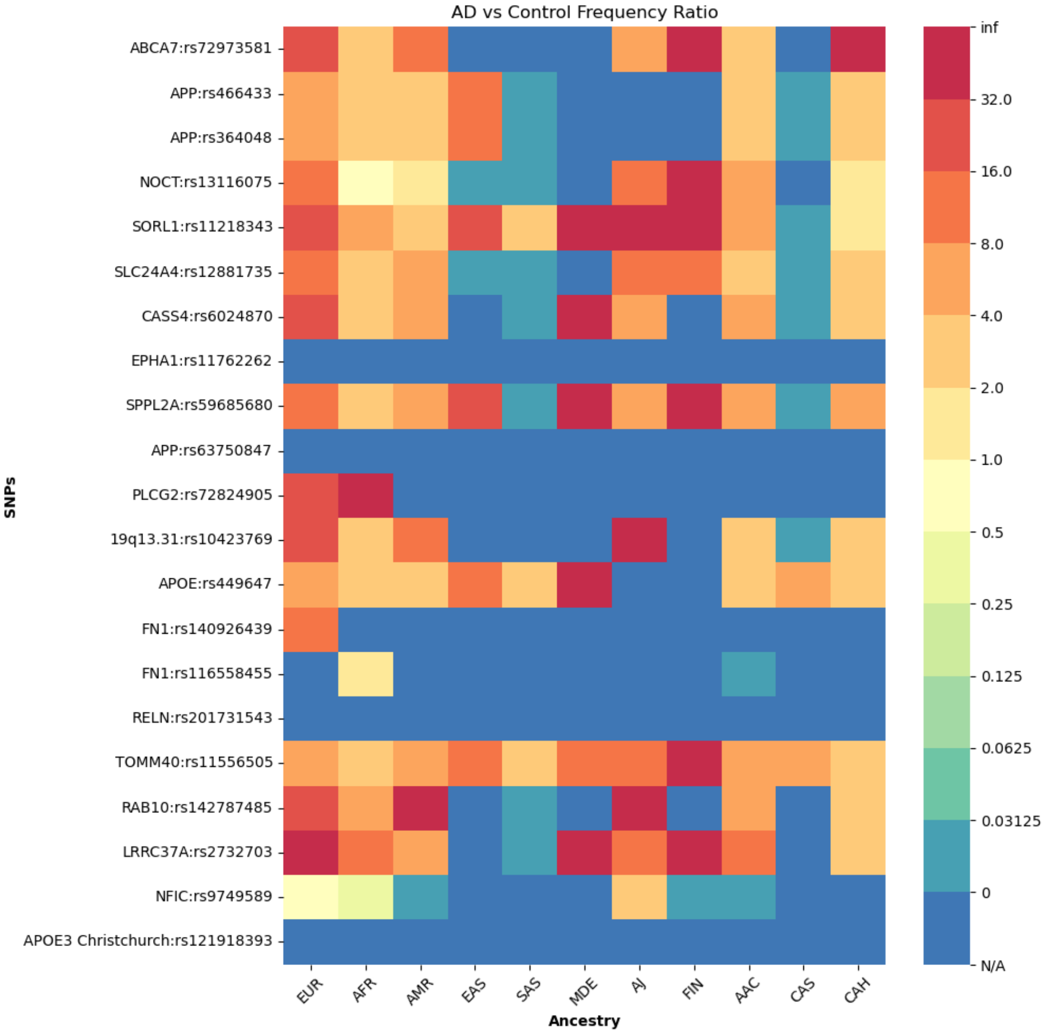

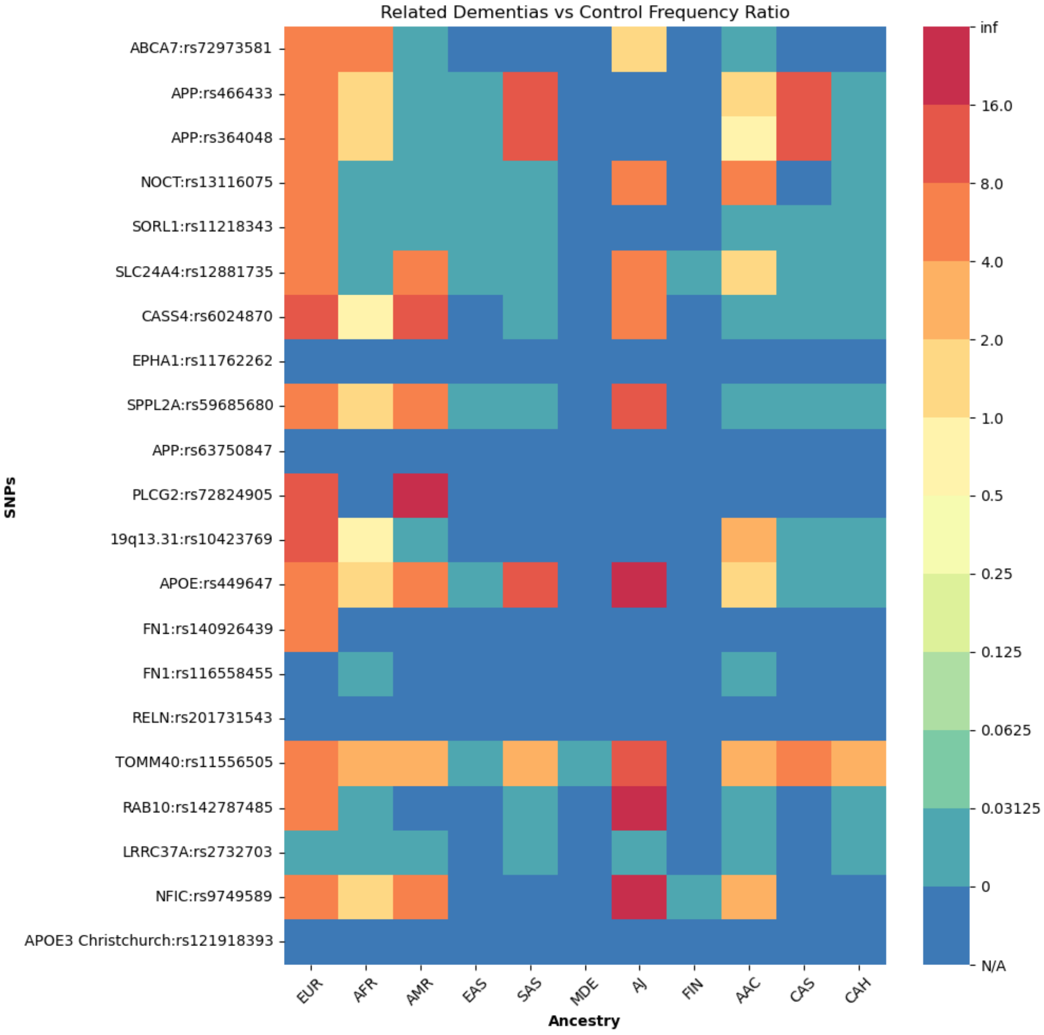

### Supplementary Figure 4C

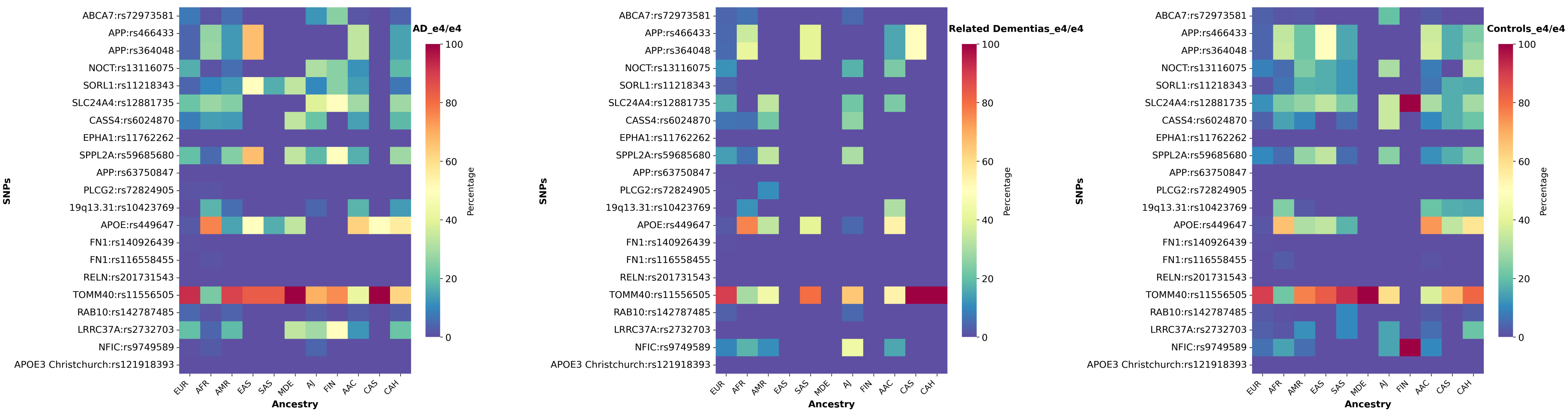

### Supplementary Figure 4D

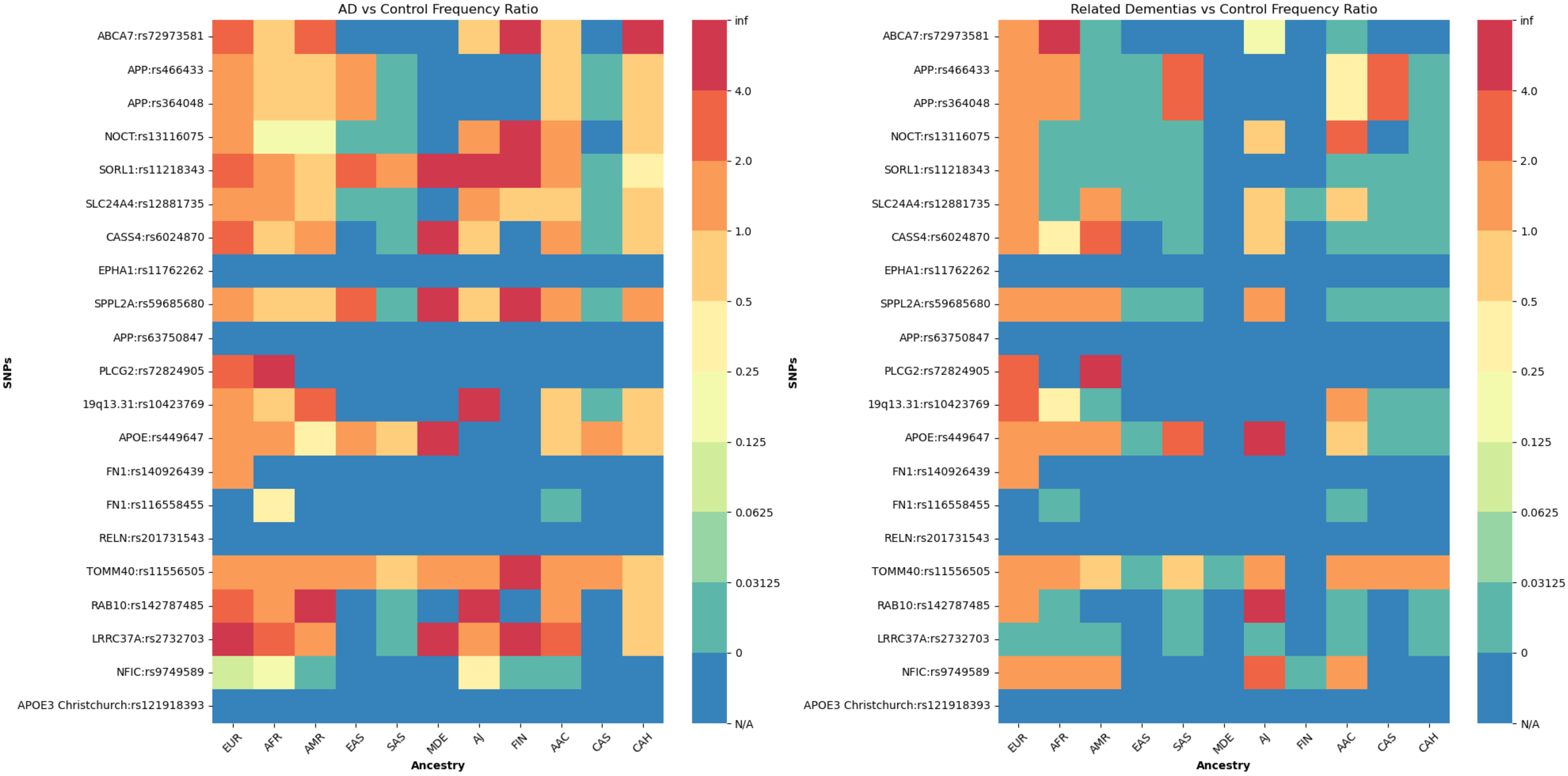
